# Human Leukocyte Antigen Homozygosity Contributes to Sensitization in Kidney Transplant Candidates

**DOI:** 10.1101/2022.01.13.22269268

**Authors:** Joshua A. Rushakoff, Loren Gragert, Marcelo J Pando, Darren Stewart, Edmund Huang, Irene Kim, Stanley Jordan, Kelsi Lindblad, Xiaohai Zhang, Peter Lalli, Jignesh K. Patel, Jon A. Kobashigawa, Evan P. Kransdorf

## Abstract

**Background:** Homozygosity for human leukocyte antigens (HLA) has been associated with adverse outcomes after viral infection as well as pregnancy-induced HLA sensitization. We sought to assess the relationship between HLA locus homozygosity and the level of HLA antibody sensitization.

**Methods:** We measured sensitization using the calculated panel reactive antibody (CPRA) value for a large cohort of 147,461 patients added to the US OPTN/UNOS kidney transplant waitlist between December 2014 and December 2019. We used multinomial logistic modeling to compare 62,510 sensitized patients to 84,955 unsensitized controls.

**Results:** We found that the number of homozygous HLA loci was strongly associated with the level of sensitization. Within highly- or extremely-sensitized candidates, women displayed a higher relative abundance of HLA homozygosity at multiple HLA loci as compared to men, with attenuation of this effect in Black candidates. In multinomial logistic modeling, the number of homozygous HLA loci was an independent predictor of sensitization and interacted with female sex but not with other factors associated with sensitization.

**Conclusions:** This study shows that HLA homozygosity is an innate genetic factor that contributes to HLA sensitization, and enhances the effect of pregnancy-related sensitization.

## INTRODUCTION

HLA sensitization remains an important disparity which limits access to deceased and living donor kidney transplant ^1,2^. Sensitization results from exposure to non-self HLA antigens, most commonly through blood transfusions ^3,4^, pregnancy ^5,6^, and previous organ transplantation ^7,8^. Patient characteristics such as ethnicity ^7,9^ are also associated with the level of sensitization, although the biological mechanisms of this remain unclear.

Several aspects of the epidemiology of sensitization remain enigmatic. First, sensitization is a pervasive problem, affecting 25-40% of candidates for solid organ ^10,11^ and stem cell transplants ^12^. Next, HLA antibodies can be identified in a small percentage of women and men apheresis donors without a history of pregnancy or transfusion ^5,13-15^ suggesting that additional factors, such as cross-reactivity/heterologous immunity, may contribute to sensitization^16,17^.

An individual’s human leukocyte antigen (HLA) genotype is comprised of two haplotypes, one inherited from each parent ^18^. Compared to other human genes, HLA loci display higher levels of heterozygosity, driven by balancing selection^19^. HLA heterozygosity has been found to be a marker of improved outcomes in a number of viral infections, including human immunodeficiency virus^20^ and hepatitis C ^21^, as well as in the response to cancer immunotherapy ^22^. Conversely, HLA homozygosity has been associated with adverse outcomes in coronavirus-19 infection^23^ and an increased risk of lymphoma development^24^. HLA homozygosity has been identified as a risk factor for pregnancy-induced HLA sensitization ^6^.

In this study, we analyze the relationship between HLA homozygosity and sensitization as measured using the calculated panel reactive antibody (CPRA) value for candidates on the kidney transplant waitlist in the United States. We hypothesized that HLA homozygosity may be an innate factor which affects the likelihood of sensitization.

## MATERIALS AND METHODS

This study was performed under an approved protocol of the Cedars-Sinai Institutional Review Board (Pro00049901).

### Study Cohort

We identified 184,828 kidney transplant candidates added to the United Network for Organ Sharing (UNOS) waitlist between December 4, 2014 (the day the Kidney Allocation System [KAS] took effect) and December 31, 2019 (Supplementary Figure 1). Because CPRA is only defined for 4 ethnic groups (White, Black, Hispanic/Latinx, and Asian), individuals of other ethnicities (n=3,459) were excluded. There were 33,904 candidates with incomplete HLA typing (of which 0.02% were missing HLA-A, 90.30% HLA-C, 0.02% HLA-B, 0.02% HLA-DR, 79.20% HLA-DQ) that were also excluded. The final study cohort comprised a non-sensitized group of 84,955 candidates and a sensitized group of 62,510 candidates.

### CPRA Calculation

CPRA was calculated for each unique set of HLA-A, -C, -B, -DR, and -DQ unacceptable-HLA antigens (UA-HLA), for each candidate. The National Marrow Donor Program (NMDP)-CPRA was used given its greater accuracy for sensitized candidates as compared to UNOS-CPRA^25^. For candidates with more than one CPRA value, the peak CPRA value prior to transplant was utilized. The CPRA calculator was developed in R^26^ and previously published^10^.

### HLA Typing and Assignment of Homozygosity

When present, we converted HLA alleles into UNOS antigen categories using a previously described mapping table^27^. We found that the frequency of homozygosity for HLA split antigens was more prevalent in each successively more sensitized group (Supplementary Table 1), suggesting that homozygosity for split antigens is functionally similar to homozygosity for broad antigens. As such, split antigens were mapped to the corresponding broad antigen using a mapping table (Supplementary Table 2). HLA homozygosity was determined based on the broad HLA antigen assignments for the HLA-A, -C, -B, -DR, and -DQ loci and candidates were designated as homozygotes if both antigens at a locus were identical. To assess if HLA antigen frequencies matched a reference population dataset, homozygosity for each antigen was calculated for each ethnic group in the study cohort and compared to the NMDP registry (Supplementary Table 3). Correlation was 0.995 for White, 0.994 for Black, 0.991 for Hispanic/Latinx and 0.979 for Asian candidates. Frequencies of HLA homozygosity for each locus in the study cohort were also compared to the NMDP registry and differed by ≤2.5% for White, Black, and Hispanic/Latinx groups, but differed by 3.7-7.2% for Asian candidates (Supplementary Table 4).

**Table 1:** Study Cohort Characteristics by Sensitization Group

|  | Non-Sensitized | Mildly-sensitized<br>(CPRA 1-69) | Highly-sensitized<br>(CPRA 70-94) | Extremely-sensitized<br>(CPRA 95-100) |
| --- | --- | --- | --- | --- |
|  | N=8,4955 | N=39,203 | N=10,995 | N=12,311 |
| Women | 30.0% | 39.8% | 67.1% | 63.6% |
| Age | 51.8 (15.0) | 51.3 (14.4) | 50.7 (13.5) | 48.1 (13.4) |
| Ethnicity |  |  |  |  |
| Caucasian | 45.9% | 42.2% | 40.8% | 34.9% |
| Black | 27.4% | 34.4% | 36.5% | 42.8% |
| Hispanic/Latinx | 19.9% | 17.0% | 16.7% | 16.7% |
| Asian | 6.8% | 6.5% | 6.1% | 5.6% |
| BMI | 31.2 (47.9) | 31.8 (52.1) | 30.8 (49.3) | 29.4 (39.6) |
| Diabetes |  |  |  |  |
| Type 1 | 3.4% | 3.3% | 3.7% | 4.8% |
| Type 2 | 39.2% | 38.0% | 32.7% | 25.8% |
| Unknown Type | 0.8% | 0.8% | 0.8% | 1.2% |
| Blood Type |  |  |  |  |
| A | 33.0% | 31.2% | 31.7% | 32.0% |
| AB | 3.87% | 3.54% | 3.91% | 3.87% |
| B | 14.5% | 15.7% | 15.0% | 15.7% |
| O | 48.6% | 49.6% | 49.4% | 48.4% |
| Prior Kidney Txp | 4.6% | 8.6% | 29.4% | 55.2% |
| Days on Waitlist | 1,169 (898) | 1,272 (857) | 1,288 (950) | 1,726 (1395) |

BMI = body mass index; Txp = transplant.

**Table 2:** Study Cohort Characteristics by Total Number of Homozygous HLA Loci

| Variable | Total Number of Homozygous HLA Loci |  |  |  |  | p-value |
| --- | --- | --- | --- | --- | --- | --- |
|  | 0 | 1 | 2 | 3 | 4-5 |  |
|  | N=63,650 | N=48,503 | N=24,331 | N=7,340 | N=3,641 |  |
| Sensitization Group |  |  |  |  |  | <0.001 |
| Non-sensitized | 59.8% | 57.4% | 54.9% | 52.8% | 50.8% |  |
| Mildly-sensitized | 26.3% | 26.8% | 26.6% | 27.4% | 25.7% |  |
| Highly-sensitized | 7.13% | 7.52% | 7.90% | 8.07% | 8.1% |  |
| Extremely-sensitized | 6.75% | 8.28% | 10.6% | 11.7% | 15.4% |  |
| Women | 38.5% | 37.9% | 37.8% | 38.2% | 39.2% | 0.094 |
| Age | 51.3 (14.7) | 51.3 (14.6) | 51.1 (14.6) | 51.3 (14.8) | 51.2 (15.2) | 0.501 |
| Ethnicity |  |  |  |  |  | <0.001 |
| White | 44.8% | 42.0% | 42.0% | 43.7% | 53.6% |  |
| Black | 30.7% | 33.8% | 31.2% | 26.8% | 16.1% |  |
| Hispanic/Latinx | 19.1% | 17.9% | 18.8% | 18.8% | 18.5% |  |
| Asian | 5.4% | 6.3% | 8.0% | 10.7% | 11.8% |  |
| BMI | 31.2 (48.7) | 31.1 (47.2) | 31.2 (48.9) | 31.4 (53.4) | 30.9 (51.2) | 0.975 |
| Diabetes |  |  |  |  |  | <0.001 |
| Type 1 | 3.9% | 2.9% | 3.3% | 3.9% | 5.5% |  |
| Type 2 | 37.3% | 37.6% | 37.4% | 35.6% | 34.5% |  |
| Unknown Type | 0.8% | 0.8% | 0.9% | 0.9% | 1.20% |  |
| Blood Type |  |  |  |  |  | 0.117 |
| A | 32.6% | 32.2% | 31.9% | 32.0% | 33.2% |  |
| AB | 3.74% | 3.83% | 3.78% | 4.05% | 3.5% |  |
| B | 14.7% | 15.0% | 15.5% | 14.4% | 14.9% |  |
| O | 48.9% | 49.0% | 48.7% | 49.5% | 48.5% |  |
| Prior Kidney Txp | 11.7% | 11.5% | 11.8% | 11.6% | 14.0% | <0.001 |

BMI = body mass index; Txp = transplant.

**Table 3.** Multivariable Model of Factors Associated with Sensitization Including Interactions with HLA Homozygosity

|  | Sensitization Group |  |  |  |  |  |
| --- | --- | --- | --- | --- | --- | --- |
|  | Mildly-sensitized<br>(CPRA 1-69) |  | Highly-sensitized<br>(CPRA 70-94) |  | Extremely-sensitized<br>(CPRA 95-100) |  |
|  | OR (95% CI) | p-value | OR (95% CI) | p-value | OR (95% CI) | p-value |
| Number of Homozygous HLA Loci | 1.08 (1.03-1.14) | 0.002 | 0.96 (0.87-1.05) | 0.383 | 1.10 (1.00-1.20) | 0.044 |
| Sex |  |  |  |  |  |  |
| Male | reference |  | reference |  | reference |  |
| Female | 1.61 (1.56-1.67) | <0.001 | 5.55 (5.22-5.90) | <0.001 | 5.36 (5.02-5.72) | <0.001 |
| Sex * HLA Homozygosity |  |  |  |  |  |  |
| Female | 0.98 (0.96-1.01) | 0.149 | 1.07 (1.02-1.11) | 0.004 | 1.15 (1.11-1.21) | <0.001 |
| Age (per decade) | 0.99 (0.98-1.00) | 0.147 | 1.01 (0.99-1.03) | 0.286 | 0.93 (0.91-0.95) | <0.001 |
| Age * HLA Homozygosity | 1.00 (0.99-1.01) | 0.897 | 1.02 (1.01-1.04) | 0.002 | 1.03 (1.02-1.05) | <0.001 |
| Ethnicity |  |  |  |  |  |  |
| White | reference |  | reference |  | reference |  |
| Black | 1.38 (1.33-1.44) | <0.001 | 1.75 (1.64-1.87) | <0.001 | 2.96 (2.76-3.19) | <0.001 |
| Hispanic/Latinx | 0.95 (0.91-1.00) | 0.050 | 1.12 (1.03-1.21) | 0.009 | 1.56 (1.42-1.70) | <0.001 |
| Asian | 0.98 (0.91-1.05) | 0.582 | 0.93 (0.82-1.06) | 0.308 | 1.14 (0.99-1.32) | 0.066 |
| Ethnicity * HLA Homozygosity |  |  |  |  |  |  |
| Black | 0.99 (0.96-1.02) | 0.450 | 0.98 (0.94-1.03) | 0.524 | 0.97 (0.92-1.02) | 0.204 |
| Hispanic/Latinx | 0.98 (0.94-1.01) | 0.179 | 1.01 (0.95-1.07) | 0.807 | 0.98 (0.93-1.04) | 0.549 |
| Asian | 1.03 (0.98-1.08) | 0.197 | 1.09 (1.01-1.178) | 0.030 | 1.05 (0.97-1.13) | 0.260 |
| Diabetes |  |  |  |  |  |  |
| None | reference |  | reference |  | reference |  |
| Type 1 | 0.95 (0.87-1.04) | 0.283 | 0.74 (0.64-0.86) | <0.001 | 0.75 (0.64-0.87) | <0.001 |
| Type 2 | 1.01 (0.98-1.05) | 0.450 | 0.97 (0.97-1.04) | 0.400 | 0.92 (0.86-0.99) | 0.031 |
| Unknown Type | 0.95 (0.79-1.15) | 0.618 | 0.84 (0.61-1.16) | 0.293 | 0.98 (0.72-1.32) | 0.871 |
| Diabetes * HLA Homozygosity |  |  |  |  |  |  |
| Type 1 | 0.98 (0.92-1.05) | 0.577 | 1.03 (0.93-1.13) | 0.589 | 1.07 (0.98-1.17) | 0.147 |
| Type 2 | 1.00 (0.98-1.03) | 0.885 | 1.01 (0.96-1.05) | 0.768 | 0.98 (0.93-1.02) | 0.328 |
| Unknown Type | 0.90 (0.79-1.03) | 0.138 | 0.89 (0.71-1.12) | 0.316 | 0.94 (0.77-1.14) | 0.543 |
| Blood Type |  |  |  |  |  |  |
| O | reference |  | reference |  | reference |  |
| A | 0.94 (0.91-0.98) | 0.001 | 0.93 (0.88-1.00) | 0.036 | 0.95 (0.89-1.02) | 0.157 |
| B | 1.04 (0.99-1.09) | 0.145 | 1.02 (0.94-1.11) | 0.652 | 1.12 (1.03-1.23) | 0.010 |
| AB | 0.88 (0.81-0.96) | 0.004 | 0.87 (0.75-1.01) | 0.066 | 0.85 (0.72-1.00) | 0.055 |
| Blood Type * HLA Homozygosity |  |  |  |  |  |  |
| A | 1.00 (0.97-1.03) | 0.868 | 1.01 (0.97-1.06) | 0.574 | 1.03 (0.98-1.07) | 0.242 |
| B | 0.99 (0.96-1.03) | 0.625 | 0.99 (0.93-1.05) | 0.685 | 0.96 (0.91-1.02) | 0.173 |
| AB | 0.98 (0.92-1.05) | 0.564 | 1.05 (0.94-1.16) | 0.380 | 1.02 (0.92-1.14) | 0.712 |
| Prior Kidney Txp |  |  |  |  |  |  |
| No | reference |  | reference |  | reference |  |
| Yes | 2.30 (2.16-2.45) | <0.001 | 12.64 (11.72-13.63) | <0.001 | 40.88 (37.95-44.03) | <0.001 |
| Prior Kidney Txp * HLA homozygosity |  |  |  |  |  |  |
| Yes | 0.93 (0.89-0.98) | 0.007 | 1.01 (0.96-1.07) | 0.682 | 0.98 (0.93-1.03) | 0.459 |

Txp = Transplant

### Statistical Analysis

The relationship between the total number of homozygous HLA loci and CPRA for each candidate was non-linear (Supplementary Figure 2). Thus, multinomial logistic modeling was used for analysis, with sensitization stratified into 4 groups based on the peak CPRA during listing/prior to transplant: (1) non-sensitized (no UA-HLA or CPRA=0), (2) mildly-sensitized (CPRA 1-69), (3) highly-sensitized (CPRA 70-94), and (4) extremely-sensitized (CPRA 95-100). CPRA thresholds for each sensitization group were based on assigning the 17 CPRA categories for candidates in the Kidney Allocation System into 3 groups of successively higher levels of sensitization composed of 6, 5, and 6 categories, respectively.

Single and multivariable multinomial modeling was performed to determine the relationship between the number of homozygous HLA loci, covariates, and sensitization. Covariates included recipient sex, age, ethnicity, diabetes mellitus, blood type, and prior kidney transplant. Interaction terms between each covariate and the total number of homozygous HLA loci were also included. For the single and multivariable models, we only considered a covariate significantly associated with sensitization if the p-value < 0.05 for all sensitization groups (*i*.*e*. mildly-, highly- and extremely-sensitized groups).

All analyses were performed with R version 4.0.5 ^26^.

## RESULTS

### Characteristics of the Study Cohort

We identified 147,465 kidney transplant candidates between 2014 and 2019 that met the inclusion criteria. This included 84,955 (57.6%) in the non-sensitized group (CPRA 0), 39,203 (26.6%) in the mildly-sensitized group (CPRA 1-69), 10,995 (7.5%) in the highly-sensitized group (CPRA 70-94), and 12,311 (8.3%) in the extremely-sensitized group (CPRA 95-100). As expected, women were present at higher percentages in the highly-and extremely-sensitized groups (Table 1). The percentage of Black candidates, the number of previous kidney transplants, and time on the waitlist increased progressively in each successively more sensitized group (p < 0.001).

### Single Locus HLA Homozygosity and Sensitization

Given homozygosity at a single HLA locus (HLA-A, -C, -B, -DR, or -DQ), we calculated the odds of presence in the mildly-, highly-, or extremely-sensitized groups (as compared to the non-sensitized group) using multinomial logistic modeling. For most HLA loci and within most ethnic groups, we found that homozygotes had significantly higher odds of being present in the mildly-sensitized, highly-sensitized, and extremely-sensitized groups (Figure 1, Supplementary Table 5). The magnitude of this homozygosity association increased for each successively more sensitized group and tended to be stronger for homozygosity at HLA-B and -DR than at HLA-A, - C, and -DQ.

**Figure 1.**
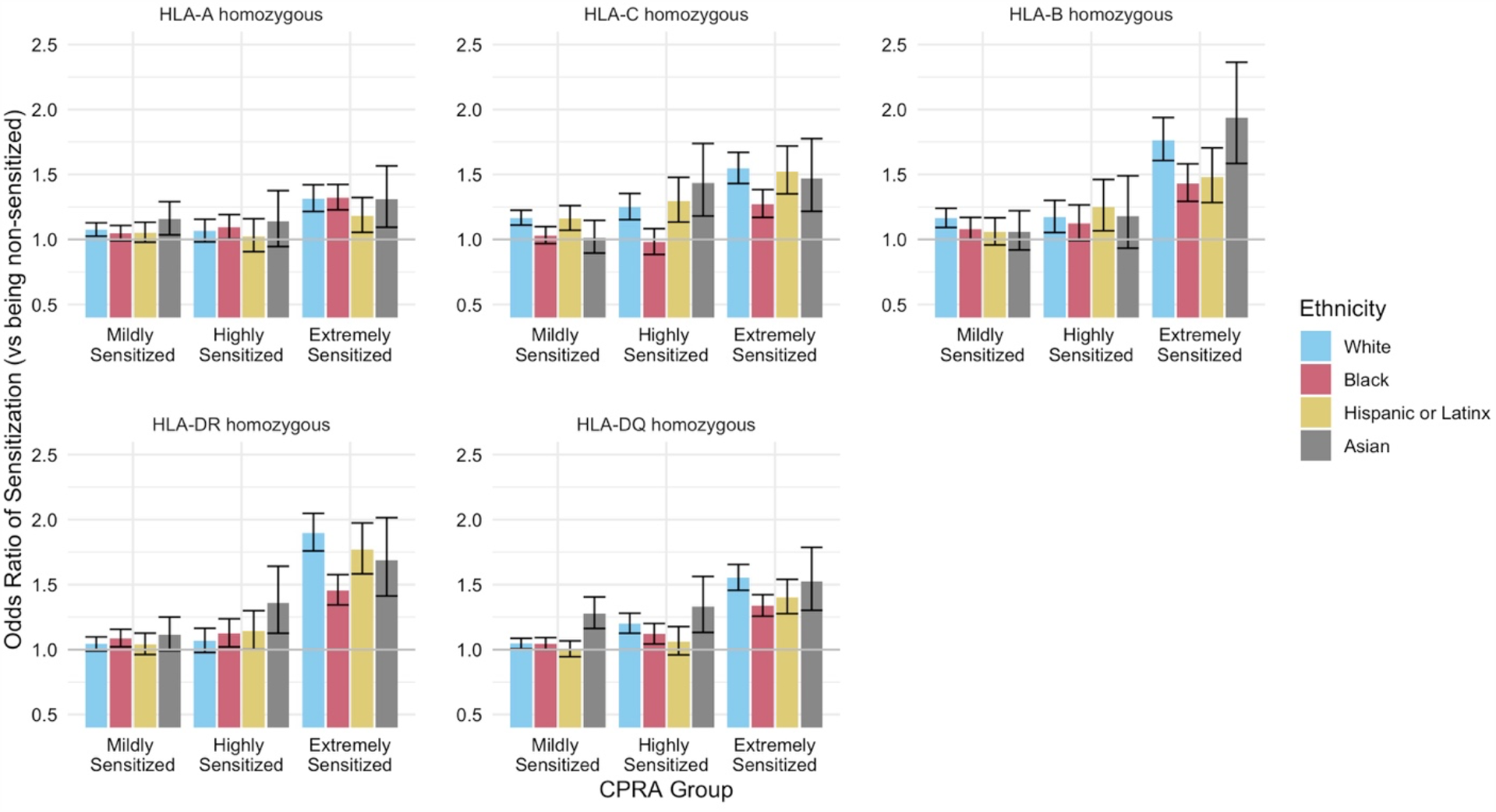
Single locus HLA homozygosity and HLA sensitization. Using multinomial logistic modeling, we calculated the odds of presence in the mildly- (CPRA 1-69), highly- (CPRA 70-95), or extremely-sensitized groups (CPRA 95-100) as compared to the non-sensitized group (no unacceptable HLA antigens of CPRA 0) given homozygosity at a single HLA locus (HLA-A, -C, -B, - DR, or -DQ). We found that homozygotes had significantly higher odds of being present in the mildly-sensitized, highly-sensitized, and extremely-sensitized groups for most HLA loci and ethnic groups.

### Multiple Homozygous HLA Loci and Sensitization

The median number of homozygous HLA loci in the study cohort was 1. Most candidates in the study cohort (83,814 or 56.8%) exhibited homozygosity at 1 or more HLA loci, and homozygosity for 2 or more loci was frequent (35,312 or 23.9%) (Table 2). The percentage of Black candidates decreased from the 0 homozygous loci group to the 5 homozygous loci group while the percentage of White and Asian candidates increased. The percentage of candidates with type 1 diabetes mellitus or a previous kidney transplant increased with a progressively higher number of homozygous HLA loci. Interestingly, the percentage of candidates in the highly-sensitized and extremely-sensitized groups generally increased with a progressively higher number of homozygous HLA loci. Other demographic factors were similar between candidates grouped by the number of homozygous HLA loci.

We further hypothesized that candidates having multiple homozygous HLA loci would have an increased risk of sensitization compared to those with a single homozygous HLA locus. Given the total number of homozygous HLA, we calculated the odds of presence in the mildly-, highly-, or extremely-sensitized groups (as compared to the non-sensitized group) using multinomial logistic modeling. Generally we found that homozygotes at multiple HLA loci had significantly higher odds of being present in the mildly-sensitized, highly-sensitized, and extremely-sensitized groups (Figure 2, Supplementary Table 6). There were exceptions to this observation: (1) Whites with HLA homozygosity at 4/5 loci were not at a higher odds of presence in the highly-sensitized group, (2) Blacks with HLA homozygosity at 4/5 loci in the mildly- and highly-sensitized groups, (3) Hispanic/Latinx with HLA homozygosity at 2, or 4/5 loci in the mildly- and highly-sensitized groups, and (4) Asians with HLA homozygosity at 3 or 4/5 loci in the mildly-sensitized groups. The magnitude of the effect tended to increase with the number of homozygous loci. For the extremely-sensitized group, there was a trend towards a decreased magnitude of the effect of homozygosity at multiple HLA loci on sensitization in Black candidates.

**Figure 2.**
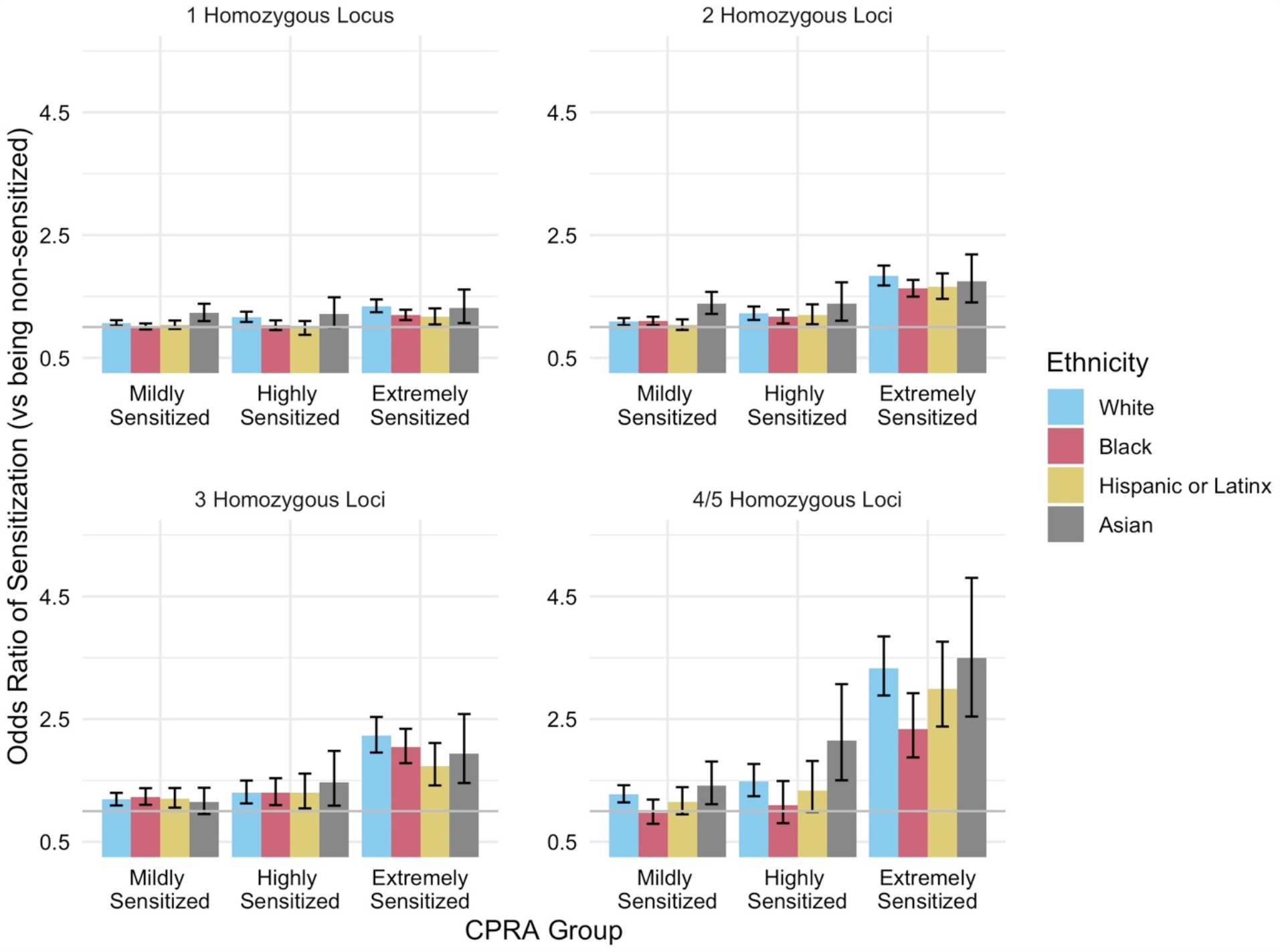
Multiple homozygous HLA Loci and HLA sensitization. Using multinomial logistic modeling, we calculated the odds of presence in the mildly- (CPRA 1-69), highly- (CPRA 70-95), or extremely-sensitized groups (CPRA 95-100) as compared to the non-sensitized group (no unacceptable HLA antigens of CPRA 0) given homozygosity at multiple HLA loci. Generally we found that homozygotes at multiple HLA loci had significantly higher odds of being present in the mildly-sensitized, highly-sensitized, and extremely-sensitized groups, with the magnitude of the effect tending to increase with the number of homozygous loci.

### Multiple Homozygous HLA Loci and Sensitization by Sex and Ethnicity

Women comprised a minority of the candidates in the study cohort (56,291 women, 38.2%) but a majority of the candidates in the highly- and extremely-sensitized groups (15,211 women, 65.2%). When assessed by ethnicity, Black women (Figure 3A) and Black men (Figure 3B) showed a higher percentage of candidates in the highly- and extremely-sensitized groups as compared to the other ethnic groups.

**Figure 3.**
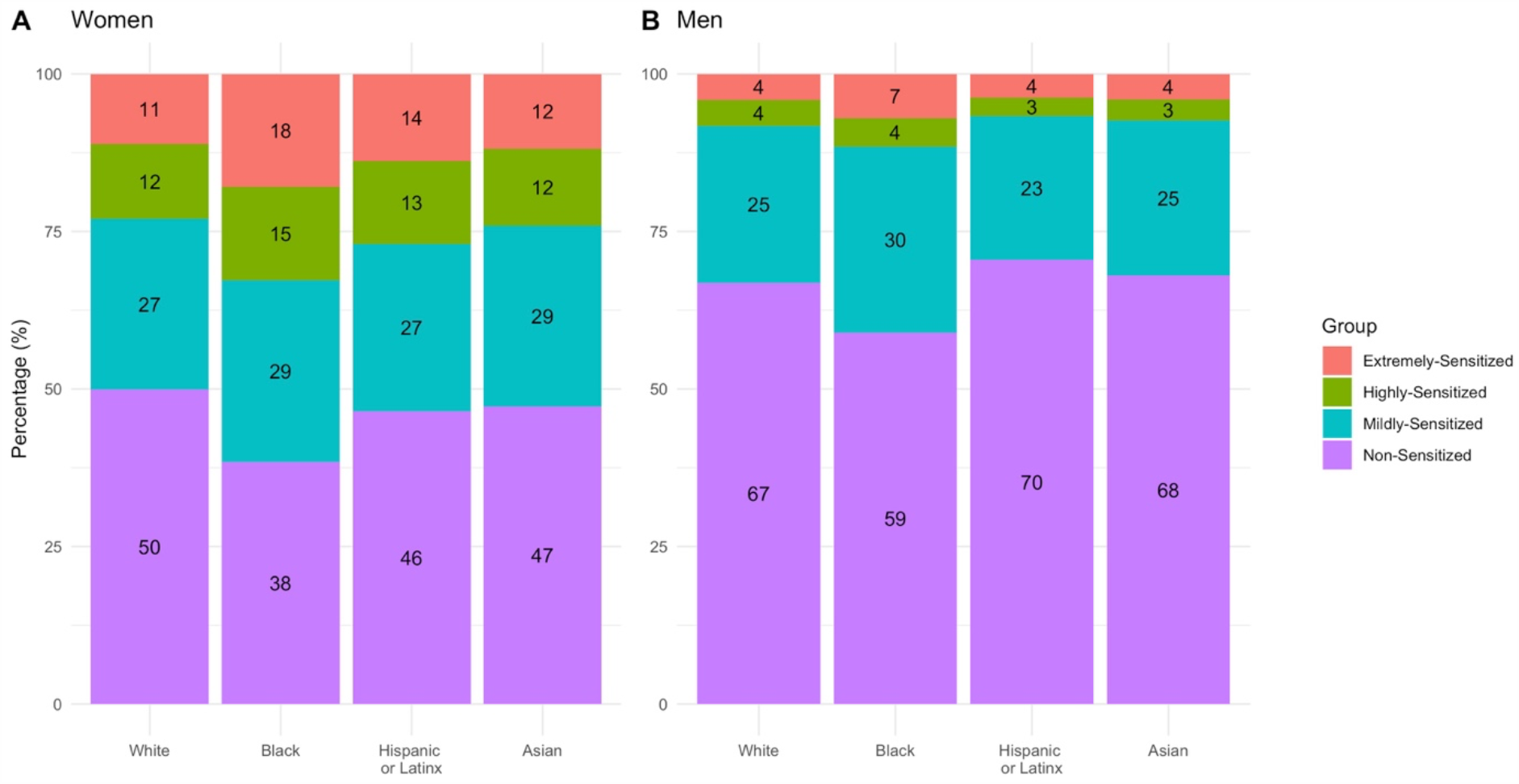
Sensitization group by sex and ethnicity. Percentage of women (A) and men (B) in each sensitization group by ethnic group. There were more Black women and men in the highly- and extremely-sensitized groups as compared to the other ethnic groups.

The study cohort was further divided into those with 0 to 1 homozygous HLA loci (112,152 or 76.1%) and those with 2 to 5 homozygous HLA loci (35,312 or 23.9%). We assessed the proportion of individuals with 2 to 5 homozygous HLA loci in each sensitization group by sex and ethnicity. For both women and men (Figure 4), there was a higher frequency of candidates with 2-5 homozygous loci in the mildly-, highly-, and extremely-sensitized groups as compared to candidates with 0 to 1 homozygous HLA loci. We found that the number of candidates with 2-5 homozygous HLA loci in the highly- and extremely-sensitized groups differed by ethnicity for both sexes (p < 0.001). Among highly- or extremely-sensitized candidates, women displayed a higher relative abundance of homozygotes at multiple HLA loci as compared to men, with attenuation of this effect in Black candidates. For women, the relative abundance of candidates in the highly- and extremely-sensitized groups (as compared to the non- and mildly-sensitized groups) with 2-5 homozygous HLA loci was highest for White and Hispanic/Latinx candidates (1.40-fold), intermediate for Asian (1.37-fold), and lowest for Black candidates (1.33-fold). Men displayed an overall lower relative abundance of candidates in the highly- and extremely- sensitized groups (as compared to the non- and mildly-sensitized groups) with 2-5 homozygous HLA loci (White: 1.23-fold, Black: 1.22-fold, Hispanic/Latinx: 1.29-fold, Asian: 1.15-fold).

**Figure 4.**
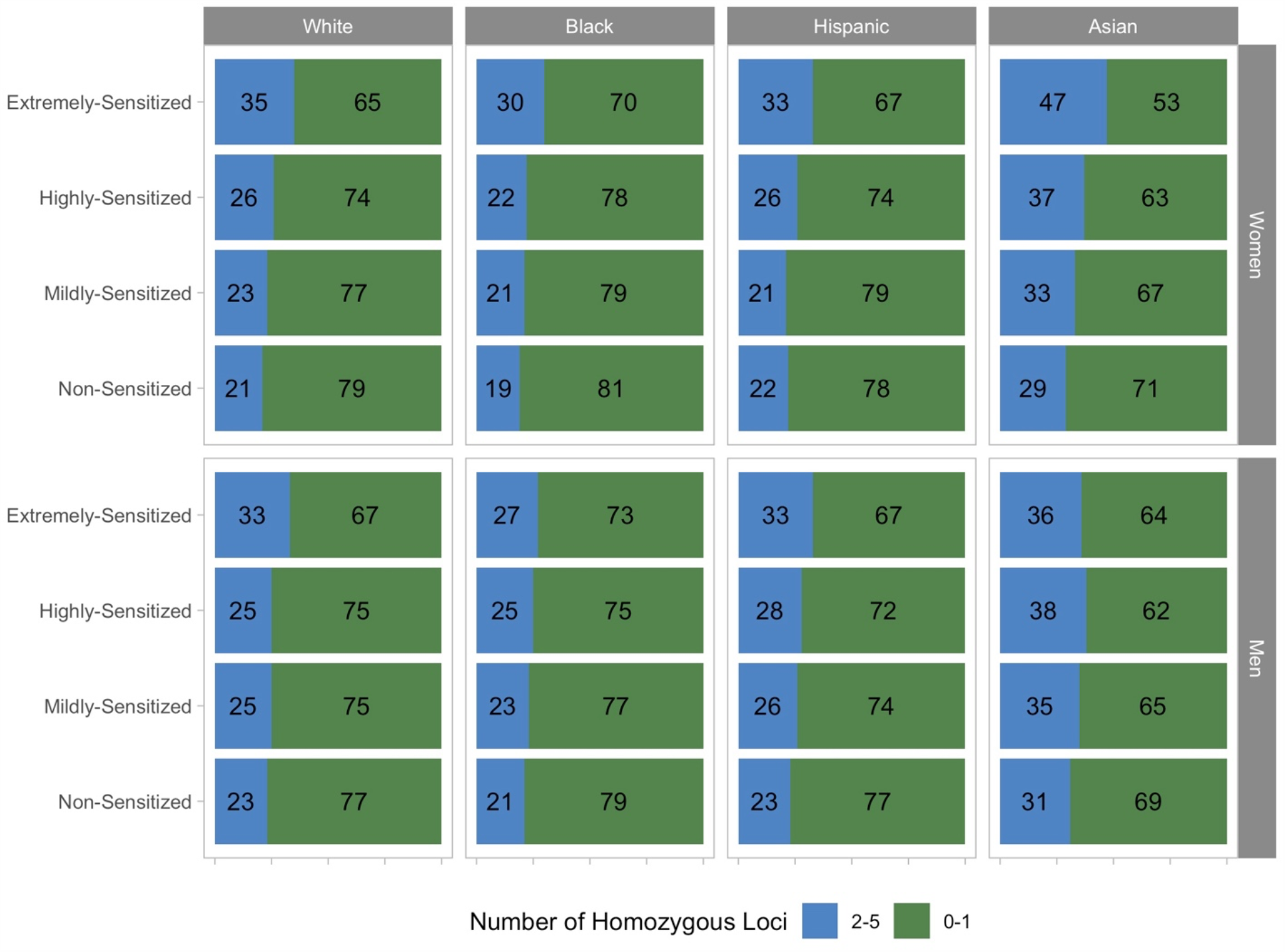
Sensitization group by HLA homozygosity and ethnicity for women and men. The cohort was further divided into those with 0 to 1 homozygous HLA loci and those with multiple (2 to 5) homozygous HLA loci. There was a higher frequency of candidates with multiple homozygous HLA loci in the mildly-, highly-, and extremely-sensitized groups as compared to candidates with 0 to 1 homozygous HLA loci for women.

### Factors Associated with Sensitization

Based on the preceding analyses, we sought to identify factors independently associated with sensitization. Covariates were analyzed in single variable multinomial models. We also included an interaction between each covariate and the total number of homozygous HLA loci. We found that the total number of homozygous HLA loci, female sex, Black ethnicity, and a history of prior kidney transplant were all associated with an increased odds of presence in the mildly-, highly-, and extremely-sensitized groups as compared to the non-sensitized group (Supplementary Table 7). Female sex, Black ethnicity, and a history of prior kidney also displayed a significant positive interaction with the total number of homozygous HLA loci. In contrast, increasing age and diabetes mellitus type 2 were associated with a decreased odds of presence in the mildly-, highly-, and extremely-sensitized groups. Increasing age displayed a significant positive interaction with the total number of homozygous HLA loci.

A multivariable model was then constructed using these factors, including their interaction, with the total number of homozygous HLA loci. We found that the total number of homozygous HLA loci remained significantly associated with an increased odds of presence in only the mildly-sensitized and extremely-sensitized groups (with the latter association of borderline significance with p=0.044; Table 3). Female sex, Black ethnicity, and a history of prior kidney transplant remained associated with an increased odds of presence in each progressively more sensitized group. Of these, female sex was the only factor displaying a significant interaction with HLA homozygosity, such that increasing number of homozygous HLA loci was associated with an increased odds of presence in the highly- (OR 1.07/homozygous locus, 95% CI 1.02-1.11, p=0.004) and extremely-sensitized groups (OR 1.15/ homozygous locus, 95% CI 1.11-1.21, p<0.001).

## DISCUSSION

In this study, we assessed the association between HLA homozygosity and sensitization. We found that homozygosity at HLA-A, -C, -B, -DR, and -DQ was associated with sensitization, and the magnitude of this effect varied by the specific locus. More significantly, we found that as the total number of homozygous loci increased, the odds of being sensitized, especially highly-sensitized or extremely-sensitized, increased. The magnitude of this effect varied by ethnicity, with a lower magnitude of this effect in Black candidates. Women comprised a majority of the highly- and extremely-sensitized candidates and we observed a higher relative abundance of women candidates with homozygosity at multiple HLA loci as compared to men. Taken together, these data show that HLA homozygosity contributes to sensitization in kidney transplant candidates.

In studies of apheresis donors, only a small percentage (<10%) of women without a history of pregnancy are sensitized ^5,13,14^. End-stage renal disease has also not been associated with sensitization in the absence of other sensitizing events ^28^. In this study we have used female sex as a surrogate marker of previous pregnancy, as parity is not available for transplant candidates in the UNOS dataset. We found that the total number of homozygous HLA loci interacted with female sex to increase the odds of sensitization, but did not interact with other factors including ethnicity and a history of prior kidney transplantation. Thus, HLA homozygosity is an innate factor that enhances the effect of pregnancy-related sensitization and contributes to the high sensitization levels for women candidates on the kidney transplant waitlist.

Although transplant rates for sensitized candidates have improved under KAS^1^, the transplant offer rate remains inversely correlated with CPRA ^25^. Further refinements to the CPRA metric, such as the inclusion of the HLA-DQA, -DPA, and -DPB loci^29^ as well as HLA alleles, may help improve access to transplant for highly sensitized women candidates. Additional studies incorporating granular data on parity may help determine if HLA homozygosity contributes to disparities in transplant access independent of CPRA.

We found that highly- and extremely-sensitized Black candidates, who comprised a larger percentage of these groups compared to other ethnicities, were enriched for candidates possessing multiple homozygous HLA loci at a lower magnitude compared to White, Hispanic/Latinx, and Asian candidates. HLA haplotypes are strongly related to ethnicity and the Black population is well known to have higher HLA heterozygosity^30^. Although we observed a strong relationship between Black ethnicity and an increased odds of presence in each progressively more sensitized group, ethnicity did not interact with the total number of homozygous HLA loci. These findings suggest that HLA homozygosity is not a mediator of the increased burden of sensitization faced by Black candidates.

The biological mechanisms responsible for the higher burden of HLA sensitization in Black kidney transplant candidates remain unclear. Differences in B cell subsets and B cell receptor (BCR) signaling have been demonstrated in healthy lymphocyte donors as well as patients with systemic lupus erythematosus of African American ancestry ^31,32^. Importantly, polymorphism at the immunoglobulin heavy chain locus is extreme^33^ and may also contribute to ethnicity-specific differences in humoral response, as has been noted after influenza vaccination ^34^. Given that highly- and extremely-sensitized Black candidates have diminished access to kidney transplant as compared to similarly sensitized White candidates^35^, further research is urgently needed to define the biologic mechanisms of sensitization in Black candidates.

The mechanism by which HLA homozygosity promotes sensitization is not established. During B cell development, V/D/J and V/J genes are randomly recombined to generate the heavy and light chains of the BCR, respectively ^36^. As a result of this stochastic process, autoreactivity of the BCR is common ^37^. Mechanisms for tolerizing autoreactive B cells include receptor editing, induction of anergy, and clonal deletion ^38^. Receptor editing^39^, the process by which the light chain gene undergoes a secondary V/J recombination and/or the heavy chain gene undergoes replacement, seems to be the major mechanism of tolerance for membrane proteins such as HLA ^40,41^.

We hypothesize that during B cell development in the bone marrow, increased HLA homozygosity leads to a less diverse repertoire of self-HLA on the cell surface, which in turn leads to less “pruning” of B cells with HLA-specific B cell receptors. Pruning of B cells is likely to occur via receptor editing. This results in a larger number and/or breadth of HLA-specific B cells, and a downstream larger number and/or breadth of HLA antibodies (Figure 5). In this way, sensitization related to HLA homozygosity is not due to a defect in tolerance *per se*, but a “loophole” in tolerance attributable to a limited repertoire of self-HLA epitopes. In support of this hypothesis, patients with systemic lupus erythematosus, which is characterized by pathologically decreased BCR editing ^42^, have been found to have elevated levels of HLA antibodies^43,44^. Further studies are needed to verify this hypothesis.

**Figure 5.**
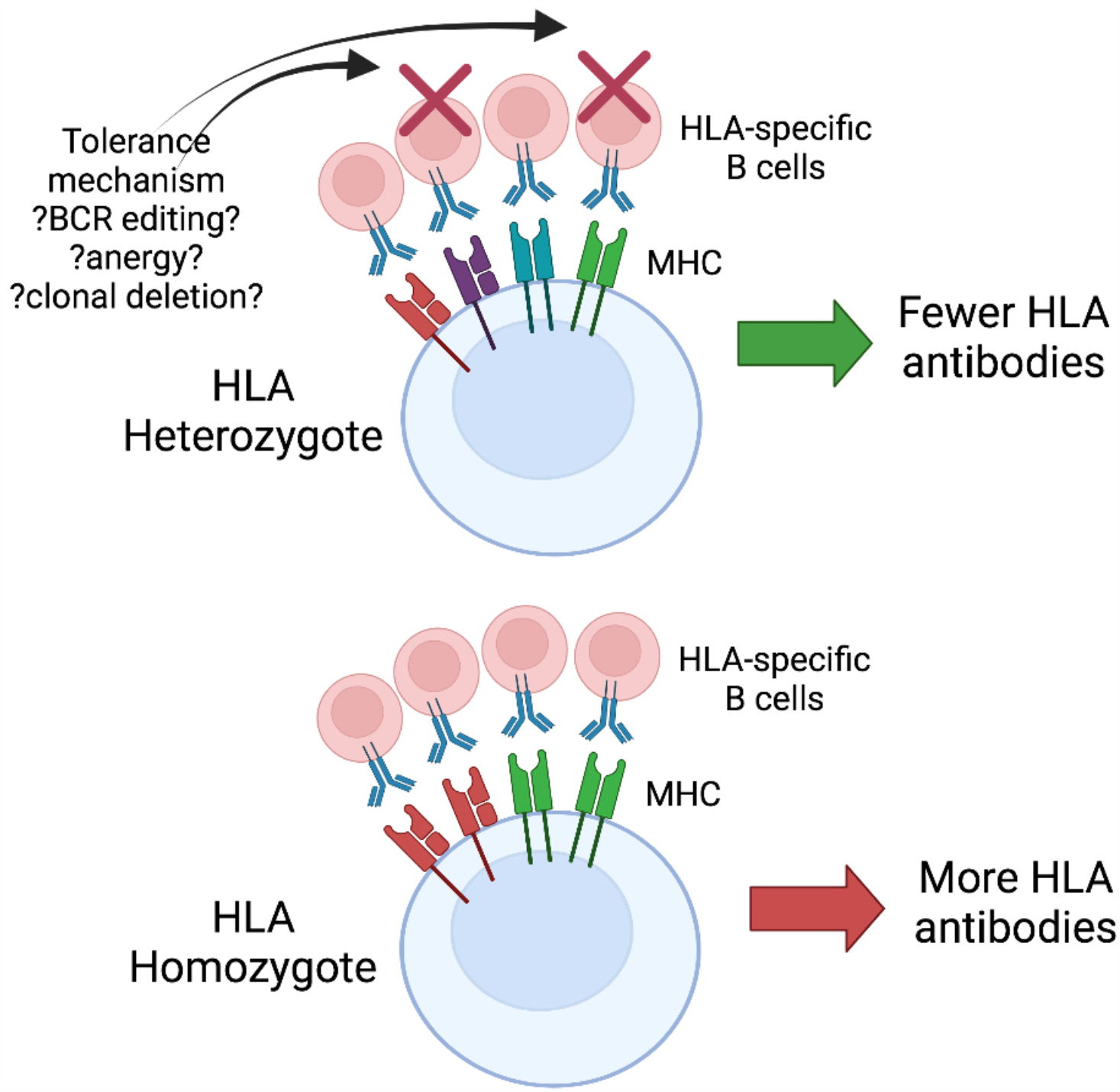
Proposed model for the mechanism of HLA homozygosity mediated sensitization. We hypothesize that increased HLA homozygosity leads to a less diverse repertoire of self-HLA on the cell surface, which in turn leads to less “pruning” of B cells with HLA-specific receptors, ultimately resulting in a larger number or breadth of HLA antibodies.

To the best of our knowledge, HLA homozygosity has not previously been identified as a factor contributing to sensitization in solid organ transplant candidates. Because HLA genes are highly polymorphic^18^, homozygosity occurs at a frequency of 10-30% for each locus. Thus, a large cohort is needed to observe the relationship between HLA homozygosity and sensitization, especially when considering candidates who have homozygosity at multiple HLA loci. Our use of a cohort of 147,461 patients facilitated our ability to identify the associations reported here.

This study has several limitations which merit discussion. There were a significant number of candidates that were excluded due to missing typing at the HLA-C and -DQ loci. Omission of these candidates could lead to skew in the cohort. Reassuringly, we found that the frequency of HLA homozygosity was similar in our cohort as compared to NMDP for Whites, Blacks, and Hispanics. We did find a higher percentage of homozygotes at all HLA loci in Asian candidates in UNOS as compared to NMDP. This could be due to differences in split versus broad antigen reporting, as Asian candidates displayed a higher percentage of homozygosity related to split antigens (Supplementary Table 1), or could be due to differences between these databases in population substructure. We focused on HLA-A, -C, -B, -DR, and -DQ, and did not investigate the role of the HLA-DP locus. Our analysis was based on low resolution (antigen level) typing. Assessing candidate HLA typing at the allelic level could allow for characterization of the effect of inter-allelic distance on sensitization as a continuous rather than dichotomized (*e*.*g*. homozygous versus heterozygous) quantity. Additionally, single antigen bead assays may miss rare specificities that are not present on the beads^45^. Granular data on the stimuli for sensitization, for example parity and history of transfusion, were not available in the database. Such data could lead to an improved understanding of how alloimmune stimuli lead to the development of HLA antibodies in the context of HLA homozygosity. HLA antibody profiles change over time and our use of maximum CPRA during listing may introduce bias. Lastly, there was no information on the use of pre-transplant desensitization therapy, which could affect antibody profiles and post-transplant outcomes.

In summary, we have found that HLA homozygosity contributes to sensitization in kidney transplant candidates. These results are particularly relevant to women as they comprise the majority of highly- and extremely-sensitized kidney transplant candidates and there is a specific interaction between the number of homozygous HLA loci and female sex, presumably via pregnancy. Thus, HLA homozygosity is an innate factor that enhances the effect of pregnancy-related sensitization and contributes to the disparity in access for sensitized women kidney transplant candidates. Further research is needed to determine the specific contribution of HLA homozygosity to differences in transplant access as well as the biologic mechanisms that underlie this effect.

## Data Availability

Data used for this study were obtained from OPTN/UNOS and are available by request to OPTN/UNOS.

## Abbreviations (alphabetical)

BCR-B: Cell Receptor
CPRA: Calculated Panel Reactive Antibody
HLA: Human Leukocyte Antigen
KAS: Kidney Allocation System
KDPI: kidney donor profile index
NMDP: National Marrow Donor Program
UA-HLA: unacceptable HLA antigens
UNOS: United Network for Organ Sharing

## ACKNOWLEDGEMENTS

This research is supported by the Smidt Heart Institute. LG is supported by the National Institute of Allergy and Infectious Disease (NIAID): U01AI152960.

## DATA AVAILABILITY STATEMENT

Data used for this study were obtained from OPTN/UNOS and are available by request to OPTN/UNOS.

## References

1. Stewart DE, Wilk AR, Toll AE, et al. Measuring and monitoring equity in access to deceased donor kidney transplantation. Am J Transplant. 2018;18(8): 1924–1935.

2. Bromberger B, Spragan D, Hashmi S, et al. Pregnancy-Induced Sensitization Promotes Sex Disparity in Living Donor Kidney Transplantation. J Am Soc Nephrol. 2017;28(10): 3025–3033.

3. Yabu JM, Anderson MW, Kim D, et al. Sensitization from transfusion in patients awaiting primary kidney transplant. Nephrol Dial Transplant. 2013;28(11): 2908–2918.

4. Leffell MS, Kim D, Vega RM, et al. Red blood cell transfusions and the risk of allosensitization in patients awaiting primary kidney transplantation. Transplantation. 2014;97(5): 525–533.

5. Triulzi DJ, Kleinman S, Kakaiya RM, et al. The effect of previous pregnancy and transfusion on HLA alloimmunization in blood donors: implications for a transfusion-related acute lung injury risk reduction strategy. Transfusion. 2009;49(9): 1825–1835.

6. Honger G, Fornaro I, Granado C, Tiercy JM, Hosli I, Schaub S. Frequency and determinants of pregnancy-induced child-specific sensitization. Am J Transplant. 2013;13(3): 746–753.

7. Meier-Kriesche HU, Scornik JC, Susskind B, Rehman S, Schold JD. A lifetime versus a graft life approach redefines the importance of HLA matching in kidney transplant patients. Transplantation. 2009;88(1): 23–29.

8. Picascia A, Grimaldi V, Sabia C, Napoli C. Comprehensive assessment of sensitizing events and anti-HLA antibody development in women awaiting kidney transplantation. Transpl Immunol. 2016;36: 14–19.

9. Baxter-Lowe LA, Kucheryavaya A, Tyan D, Reinsmoen N. CPRA for allocation of kidneys in the US: More candidates >/=98% CPRA, lower positive crossmatch rates and improved transplant rates for sensitized patients. Hum Immunol. 2016;77(5): 395–402.

10. Kransdorf EP, Kittleson MM, Patel JK, Pando MJ, Steidley DE, Kobashigawa JA. Calculated panel-reactive antibody predicts outcomes on the heart transplant waiting list. J Heart Lung Transplant. 2017;36(7): 787–796.

11. Tague LK, Witt CA, Byers DE, et al. Association between Allosensitization and Waiting List Outcomes among Adult Lung Transplant Candidates in the United States. Ann Am Thorac Soc. 2019;16(7): 846–852.

12. Huo MR, Xu YJ, Zhai SZ, et al. Prevalence and risk factors of antibodies to human leukocyte antigens in haploidentical stem cell transplantation candidates: A multi-center study. Hum Immunol. 2018;79(9): 672–677.

13. Densmore TL, Goodnough LT, Ali S, Dynis M, Chaplin H. Prevalence of HLA sensitization in female apheresis donors. Transfusion. 1999;39(1): 103–106.

14. De Clippel D, Baeten M, Torfs A, et al. Screening for HLA antibodies in plateletpheresis donors with a history of transfusion or pregnancy. Transfusion. 2014;54(12): 3036–3042.

15. Morales-Buenrostro LE, Terasaki PI, Marino-Vazquez LA, Lee JH, El-Awar N, Alberu J. “Natural” human leukocyte antigen antibodies found in nonalloimmunized healthy males. Transplantation. 2008;86(8): 1111–1115.

16. Locke JE, Zachary AA, Warren DS, et al. Proinflammatory events are associated with significant increases in breadth and strength of HLA-specific antibody. Am J Transplant. 2009;9(9): 2136–2139.

17. van den Heuvel H, Heutinck KM, van der Meer-Prins EMW, et al. Allo-HLA Cross-Reactivities of Cytomegalovirus-, Influenza-, and Varicella Zoster Virus-Specific Memory T Cells Are Shared by Different Healthy Individuals. Am J Transplant. 2017;17(8): 2033–2044.

18. Kransdorf EP, Pando MJ, Gragert L, Kaplan B. HLA Population Genetics in Solid Organ Transplantation. Transplantation. 2017;101(9): 1971–1976.

19. Solberg OD, Mack SJ, Lancaster AK, et al. Balancing selection and heterogeneity across the classical human leukocyte antigen loci: a meta-analytic review of 497 population studies. Hum Immunol. 2008;69(7): 443–464.

20. Arora J, Pierini F, McLaren PJ, Carrington M, Fellay J, Lenz TL. HLA Heterozygote Advantage against HIV-1 Is Driven by Quantitative and Qualitative Differences in HLA Allele-Specific Peptide Presentation. Mol Biol Evol. 2020;37(3): 639–650.

21. Hraber P, Kuiken C, Yusim K. Evidence for human leukocyte antigen heterozygote advantage against hepatitis C virus infection. Hepatology. 2007;46(6): 1713–1721.

22. Chowell D, Morris LGT, Grigg CM, et al. Patient HLA class I genotype influences cancer response to checkpoint blockade immunotherapy. Science. 2018;359(6375): 582–587.

23. Shkurnikov M, Nersisyan S, Jankevic T, et al. Association of HLA Class I Genotypes With Severity of Coronavirus Disease-19. Front Immunol. 2021;12: 641900.

24. Wang SS, Carrington M, Berndt SI, et al. HLA Class I and II Diversity Contributes to the Etiologic Heterogeneity of Non-Hodgkin Lymphoma Subtypes. Cancer Res. 2018;78(14): 4086–4096.

25. Kransdorf EP, Pando MJ, Stewart D, et al. Stem cell donor HLA typing improves CPRA in kidney allocation. Am J Transplant. 2021;21(1): 138–147.

26. The R Project for Statistical Computing.

27. Kaur N, Kransdorf EP, Pando MJ, et al. Mapping molecular HLA typing data to UNOS antigen equivalents. Hum Immunol. 2018;79(11): 781–789.

28. Hung SY, Lin TM, Chang MY, et al. Risk factors of sensitization to human leukocyte antigen in end-stage renal disease patients. Hum Immunol. 2014;75(6): 531–535.

29. Tinckam KJ, Liwski R, Pochinco D, et al. cPRA Increases With DQA, DPA, and DPB Unacceptable Antigens in the Canadian cPRA Calculator. Am J Transplant. 2015;15(12): 3194–3201.

30. Gragert L, Madbouly A, Freeman J, Maiers M. Six-locus high resolution HLA haplotype frequencies derived from mixed-resolution DNA typing for the entire US donor registry. Hum Immunol. 2013;74(10): 1313–1320.

31. Longo DM, Louie B, Mathi K, et al. Racial differences in B cell receptor signaling pathway activation. J Transl Med. 2012;10: 113.

32. Menard LC, Habte S, Gonsiorek W, et al. B cells from African American lupus patients exhibit an activated phenotype. JCI Insight. 2016;1(9): e87310.

33. Watson CT, Breden F. The immunoglobulin heavy chain locus: genetic variation, missing data, and implications for human disease. Genes Immun. 2012;13(5): 363–373.

34. Avnir Y, Watson CT, Glanville J, et al. IGHV1-69 polymorphism modulates anti-influenza antibody repertoires, correlates with IGHV utilization shifts and varies by ethnicity. Sci Rep. 2016;6: 20842.

35. Kulkarni S, Ladin K, Haakinson D, Greene E, Li L, Deng Y. Association of Racial Disparities With Access to Kidney Transplant After the Implementation of the New Kidney Allocation System. JAMA Surg. 2019;154(7): 618–625.

36. Jackson KJ, Kidd MJ, Wang Y, Collins AM. The shape of the lymphocyte receptor repertoire: lessons from the B cell receptor. Front Immunol. 2013;4: 263.

37. Wardemann H, Yurasov S, Schaefer A, Young JW, Meffre E, Nussenzweig MC. Predominant autoantibody production by early human B cell precursors. Science. 2003;301(5638): 1374–1377.

38. Nemazee D. Mechanisms of central tolerance for B cells. Nat Rev Immunol. 2017;17(5): 281–294.

39. Luning Prak ET, Monestier M, Eisenberg RA. B cell receptor editing in tolerance and autoimmunity. Ann N Y Acad Sci. 2011;1217: 96–121.

40. Halverson R, Torres RM, Pelanda R. Receptor editing is the main mechanism of B cell tolerance toward membrane antigens. Nat Immunol. 2004;5(6): 645–650.

41. Caucheteux SM, Vernochet C, Wantyghem J, Gendron MC, Kanellopoulos-Langevin C. Tolerance induction to self-MHC antigens in fetal and neonatal mouse B cells. Int Immunol. 2008;20(1): 11–20.

42. Panigrahi AK, Goodman NG, Eisenberg RA, Rickels MR, Naji A, Luning Prak ET. RS rearrangement frequency as a marker of receptor editing in lupus and type 1 diabetes. J Exp Med. 2008;205(13): 2985–2994.

43. Jackman RP, Cruz GI, Nititham J, et al. Increased alloreactive and autoreactive antihuman leucocyte antigen antibodies associated with systemic lupus erythematosus and rheumatoid arthritis. Lupus Sci Med. 2018;5(1): e000278.

44. Tozkir H, Pamuk ON, Duymaz J, et al. Increased frequency of class I and II anti-human leukocyte antigen antibodies in systemic lupus erythematosus and scleroderma and associated factors: a comparative study. Int J Rheum Dis. 2016;19(12): 1304–1309.

45. Jani V, Ingulli E, Mekeel K, Morris GP. Root cause analysis of limitations of virtual crossmatch for kidney allocation to highly-sensitized patients. Hum Immunol. 2017;78(2): 72–79.

