## Supplemental Data for "Human Leukocyte Antigen Homozygosity Contributes to Sensitization in Kidney Transplant Candidates"

**Supplemental Methods.**

**Supplementary Figure 1.** Cohort Selection Criteria

**Supplementary Table 1.** Percentage of Candidates with HLA Homozygosity Due to Split Antigen Homozygosity

**Supplementary Table 2.** Mapping Table for HLA Splits to Broad Antigens

**Supplementary Table 3.** HLA Frequency in the Study Cohort Compared to the NMDP Registry (By Antigen)

### **Supplementary Table 4.** Frequency of HLA Homozygosity in the Study Cohort Compared to the NMDP Registry

**Supplementary Figure 2.** Plot of CPRA Sensitization by Total Number of Homozygous Loci

**Supplementary Table 5.** Odds of Presence in Sensitization Groups Based on Homozygosity at a Single HLA Locus

**Supplementary Table 6.** Odds of Presence in Sensitization Groups Based on Homozygosity at Multiple HLA Loci

### **Supplementary Table 7.** Single Variable Modeling of Factors Associated with Sensitization

### **Supplementary Methods**

**Statistical Analysis**

Relative abundance of candidates with multiple homozygous HLA loci (MHHL) was calculated as:

$$\frac{Percent candidates with MHHL in highly- and extremely-sensitized groups}{Percent candidates with MHHL in non- and mildly-sensitized groups}$$

Continuous variables are expressed as mean +/- standard deviation or median and interquartile range. Categorical data is presented as counts and percentages. ANOVA and chi-squared tests were used to compare continuous and categorical baseline characteristics, respectively. Kruskal-Wallis test was used to compare sensitization levels across ethnicities.

We calculated the number of HLA mismatches at HLA-A, -C, -B, -DR and -DQ between transplant recipients and their donors. Mismatches were assigned based on the number of mismatched antigens for each locus. A 2-sided p-value <0.05 was considered to indicate statistical significance. All analyses were completed in R (R version 4.0.0 (2020-04-24)).

**Supplementary Figure 1:** Cohort Selection Criteria

Patients Listed for Kidney Transplant Alone
12/04/14 - 12/31/19
(n = 184,828)

Excluded (n = 37,364)

Incomplete HLA typing (n = 33,904)

Non CAU/AFA/HIS/API (n = 3,459)

Sensitized Group

(n = 62,510)

Non-sensitized Group

(n = 84,955)

We identified 184,828 patients who were added the kidney transplant waitlist during the study period. Patients with incomplete HLA typing and/or ethnicity other than White, Black, Hispanic/Latinx, or Asian were excluded. The cohort was divided into non-sensitized (n= 84,955) and sensitized (n=62,510) cohorts.

### **Supplementary Table 1.** Percentage of Candidates with HLA Homozygosity Due to Split Antigen Homozygosity

|  | No Homozygous HLA Loci | Homozygous HLA Loci,  No Split Antigens | Homozygous HLA Loci,  Split Antigens |
| --- | --- | --- | --- |
| White |  |  |  |
| Non-sensitized | 46.05 | 17.64 | 36.31 |
| Mildly-sensitized | 43.84 | 18.81 | 37.35 |
| Highly-sensitized | 41.42 | 18.14 | 40.44 |
| Extremely-sensitized | 34.07 | 20.00 | 45.92 |
| Black |  |  |  |
| Non-sensitized | 43.83 | 12.15 | 44.01 |
| Mildly-sensitized | 42.69 | 12.34 | 44.97 |
| Highly-sensitized | 41.84 | 11.24 | 46.92 |
| Extremely-sensitized | 35.89 | 13.66 | 50.45 |
| Hispanic/Latinx |  |  |  |
| Non-sensitized | 45.30 | 14.61 | 40.08 |
| Mildly-sensitized | 43.98 | 16.5 | 39.52 |
| Highly-sensitized | 43.29 | 15.01 | 41.7 |
| Extremely-sensitized | 36.70 | 15.45 | 47.86 |
| Asian |  |  |  |
| Non-sensitized | 38.57 | 11.85 | 49.58 |
| Mildly-sensitized | 32.95 | 11.55 | 55.5 |
| Highly-sensitized | 31.69 | 14.20 | 54.11 |
| Extremely-sensitized | 27.52 | 15.71 | 56.77 |

### **Supplementary Table 2.** Mapping Table for HLA Splits to Broad Antigens

| Broad | Split |
| --- | --- |
| A2 | A203, A210 |
| A9 | A23, A24, A2403 |
| A10 | A25, A26, A34, A66, A6601, A6602 |
| A19 | A29, A30, A31, A32, A33, A74 |
| A28 | A68, A69 |
| B5 | B51, B52, B5102, B5103 |
| B7 | B703 |
| B12 | B44, B45 |
| B14 | B64, B65 |
| B15 | B62, B63, B75, B76, B77 |
| B16 | B38, B39, B3901, B3902, 3905 |
| B17 | B57, B58 |
| B21 | B49, B50, B4005 |
| B22 | B54, B55, B56 |
| B27 | B2708 |
| B40 | B60, B61 |
| B70 | B71, B72 |
| B82 | B8201 |
| C03 | C09, C10 |
| DR1 | DR103 |
| DR2 | DR15, DR16 |
| DR3 | DR17, DR18 |
| DR5 | DR11, DR12 |
| DR6 | DR13, DR14, DR1403, DR1404 |
| DQ1 | DQ5, DQ6 |
| DQ3 | DQ7, DQ8, DQ9 |

### **Supplementary Table 3.** HLA Frequency in the Study Cohort Compared to the NMDP Registry (By Antigen)

|  | HLA-A Antigens | | | | | | | | | | | | | | | | | | | | | | | | | | | | | | |  | | |  | | |  | | | |
| --- | --- | --- | --- | --- | --- | --- | --- | --- | --- | --- | --- | --- | --- | --- | --- | --- | --- | --- | --- | --- | --- | --- | --- | --- | --- | --- | --- | --- | --- | --- | --- | --- | --- | --- | --- | --- | --- | --- | --- | --- | --- |
|  | 1 | | | 2 | | | 3 | | | 9 | | | 10 | | | 11 | | | 19 | | | 28 | | | 36 | | 43 | | 80 | | |  | | |  | | |  | | | |
| White |  | | |  | | |  | | |  | | |  | | |  | | |  | | |  | | |  | |  | |  | | |  | | |  | | |  | | | |
| NMDP | 2.535 | | | 8.207 | | | 2.049 | | | 1.270 | | | 0.382 | | | 0.350 | | | 1.796 | | | 0.187 | | | 0.000 | | 0.000 | | 0.000 | | |  | | |  | | |  | | | |
| Study | 2.830 | | | 8.138 | | | 1.873 | | | 1.299 | | | 0.447 | | | 0.384 | | | 2.023 | | | 0.191 | | | 0.003 | | 0.000 | | 0.002 | | |  | | |  | | |  | | | |
| Black |  | | |  | | |  | | |  | | |  | | |  | | |  | | |  | | |  | |  | |  | | |  | | |  | | |  | | | |
| NMDP | 0.259 | | | 3.308 | | | 0.712 | | | 1.840 | | | 0.589 | | | 0.020 | | | 10.415 | | | 1.011 | | | 0.064 | | 0.000 | | 0.006 | | |  | | |  | | |  | | | |
| Study | 0.278 | | | 3.287 | | | 0.603 | | | 1.854 | | | 0.599 | | | 0.033 | | | 10.199 | | | 1.012 | | | 0.093 | | 0.000 | | 0.004 | | |  | | |  | | |  | | | |
| Hispanic/Latinx |  | | |  | | |  | | |  | | |  | | |  | | |  | | |  | | |  | |  | |  | | |  | | |  | | |  | | | |
| NMDP | 0.556 | | | 7.705 | | | 0.676 | | | 2.672 | | | 0.255 | | | 0.216 | | | 4.220 | | | 0.918 | | | 0.000 | | 0.000 | | 0.000 | | |  | | |  | | |  | | | |
| Study | 0.473 | | | 8.144 | | | 0.684 | | | 2.971 | | | 0.273 | | | 0.218 | | | 4.624 | | | 1.405 | | | 0.000 | | 0.000 | | 0.004 | | |  | | |  | | |  | | | |
| Asian |  | | |  | | |  | | |  | | |  | | |  | | |  | | |  | | |  | |  | |  | | |  | | |  | | |  | | | |
| NMDP | 0.511 | | | 4.622 | | | 0.142 | | | 4.426 | | | 0.361 | | | 3.409 | | | 3.650 | | | 0.087 | | | 0.000 | | 0.000 | | 0.000 | | |  | | |  | | |  | | | |
| Study | 0.672 | | | 4.438 | | | 0.155 | | | 6.994 | | | 1.521 | | | 6.238 | | | 2.690 | | | 0.310 | | | 0.000 | | 0.000 | | 0.000 | | |  | | |  | | |  | | | |
|  | HLA-C Antigens | | | | | | | | | | | | | | | | | | | | | | | | | | | | | | | | | | | | | | | | |
|  | 1 | | 2 | | | 3 | | | 4 | | | 5 | | | 6 | | | 7 | | | 8 | | | 12 | | | | 14 | | | 15 | | | 16 | | | 17 | | | 18 | |
| White |  | |  | | |  | | |  | | |  | | |  | | |  | | |  | | |  | | | |  | | |  | | |  | | |  | | |  | |
| NMDP | 0.222 | | 0.220 | | | 1.650 | | | 1.331 | | | 0.683 | | | 0.911 | | | 10.179 | | | 0.135 | | | 0.444 | | | | 0.000 | | | 0.075 | | | 0.116 | | | 0.000 | | | 0.000 | |
| Study | 0.137 | | 0.218 | | | 1.898 | | | 1.393 | | | 0.870 | | | 0.862 | | | 9.903 | | | 0.174 | | | 0.594 | | | | 0.017 | | | 0.096 | | | 0.142 | | | 0.025 | | | 0.000 | |
| Black |  | |  | | |  | | |  | | |  | | |  | | |  | | |  | | |  | | | |  | | |  | | |  | | |  | | |  | |
| NMDP | 0.067 | | 0.794 | | | 0.774 | | | 4.191 | | | 0.113 | | | 1.398 | | | 6.960 | | | 0.203 | | | 0.026 | | | | 0.000 | | | 0.034 | | | 0.944 | | | 0.000 | | | 0.000 | |
| Study | 0.024 | | 0.881 | | | 0.788 | | | 4.244 | | | 0.067 | | | 0.879 | | | 4.201 | | | 0.287 | | | 0.037 | | | | 0.037 | | | 0.054 | | | 0.981 | | | 0.382 | | | 0.115 | |
| Hispanic/Latinx |  | |  | | |  | | |  | | |  | | |  | | |  | | |  | | |  | | | |  | | |  | | |  | | |  | | |  | |
| NMDP | 0.366 | | 0.158 | | | 1.467 | | | 2.836 | | | 0.329 | | | 0.441 | | | 6.265 | | | 0.599 | | | 0.282 | | | | 0.000 | | | 0.197 | | | 0.376 | | | 0.000 | | | 0.000 | |
| Study | 0.353 | | 0.269 | | | 1.955 | | | 3.113 | | | 0.258 | | | 0.462 | | | 5.607 | | | 0.779 | | | 0.284 | | | | 0.018 | | | 0.255 | | | 0.444 | | | 0.036 | | | 0.011 | |
| Asian |  | |  | | |  | | |  | | |  | | |  | | |  | | |  | | |  | | | |  | | |  | | |  | | |  | | |  | |
| NMDP | 2.726 | | 0.004 | | | 2.941 | | | 1.319 | | | 0.006 | | | 0.594 | | | 4.669 | | | 0.646 | | | 0.572 | | | | 0.000 | | | 0.575 | | | 0.009 | | | 0.000 | | | 0.000 | |
| Study | 1.842 | | 0.010 | | | 2.700 | | | 2.793 | | | 0.010 | | | 0.641 | | | 6.869 | | | 1.076 | | | 0.900 | | | | 0.331 | | | 0.569 | | | 0.031 | | | 0.021 | | | 0.000 | |
|  | HLA-B Antigens | | | | | | | | | | | | | | | | | | | | | | | | | | | | | | | | | | | | | | | | |
|  | 5 | 7 | | | 8 | | | 12 | | | 13 | | | 14 | | | 15 | | | 16 | | | 17 | | | 18 | | | | 21 | | | 22 | | | 27 | | | 35 | | 37 |
| White |  |  | | |  | | |  | | |  | | |  | | |  | | |  | | |  | | |  | | | |  | | |  | | |  | | |  | |  |
| NMDP | 0.431 | 1.660 | | | 1.122 | | | 1.982 | | | 0.073 | | | 0.130 | | | 0.463 | | | 0.187 | | | 0.199 | | | 0.228 | | | | 0.080 | | | 0.063 | | | 0.172 | | | 0.899 | | 0.020 |
| Study | 0.440 | 1.544 | | | 1.534 | | | 2.065 | | | 0.061 | | | 0.142 | | | 0.523 | | | 0.280 | | | 0.257 | | | 0.294 | | | | 0.160 | | | 0.047 | | | 0.190 | | | 0.951 | | 0.030 |
| Black |  |  | | |  | | |  | | |  | | |  | | |  | | |  | | |  | | |  | | | |  | | |  | | |  | | |  | |  |
| NMDP | 0.151 | 0.664 | | | 0.143 | | | 1.405 | | | 0.006 | | | 0.102 | | | 0.124 | | | 0.024 | | | 1.557 | | | 0.103 | | | | 0.136 | | | 0.004 | | | 0.016 | | | 0.531 | | 0.003 |
| Study | 0.169 | 0.671 | | | 0.176 | | | 1.550 | | | 0.015 | | | 0.089 | | | 0.139 | | | 0.028 | | | 1.522 | | | 0.096 | | | | 0.135 | | | 0.011 | | | 0.022 | | | 0.471 | | 0.011 |
| Hispanic/Latinx |  |  | | |  | | |  | | |  | | |  | | |  | | |  | | |  | | |  | | | |  | | |  | | |  | | |  | |  |
| NMDP | 0.832 | 0.401 | | | 0.174 | | | 1.425 | | | 0.016 | | | 0.261 | | | 0.288 | | | 0.717 | | | 0.159 | | | 0.172 | | | | 0.196 | | | 0.019 | | | 0.053 | | | 2.285 | | 0.006 |
| Study | 0.834 | 0.320 | | | 0.153 | | | 1.373 | | | 0.004 | | | 0.178 | | | 0.397 | | | 1.260 | | | 0.200 | | | 0.204 | | | | 0.262 | | | 0.029 | | | 0.080 | | | 2.883 | | 0.004 |
| Asian |  |  | | |  | | |  | | |  | | |  | | |  | | |  | | |  | | |  | | | |  | | |  | | |  | | |  | |  |
| NMDP | 1.335 | 0.266 | | | 0.032 | | | 0.364 | | | 0.236 | | | 0.001 | | | 1.421 | | | 0.238 | | | 0.728 | | | 0.028 | | | | 0.012 | | | 0.282 | | | 0.048 | | | 0.875 | | 0.032 |
| Study | 1.469 | 0.259 | | | 0.186 | | | 0.455 | | | 0.486 | | | 0.000 | | | 2.669 | | | 1.055 | | | 0.579 | | | 0.103 | | | | 0.041 | | | 0.435 | | | 0.083 | | | 0.848 | | 0.031 |
|  | HLA-B Antigens | | | | | | | | | | | | | | | | | | | | | | | | | | | | | | | | | | | | | | | |  |
|  | 40 | 41 | | | 42 | | | 46 | | | 47 | | | 48 | | | 53 | | | 59 | | | 67 | | | 70 | | | | 73 | | | 78 | | | 81 | | | 82 | |  |
| White |  |  | | |  | | |  | | |  | | |  | | |  | | |  | | |  | | |  | | | |  | | |  | | |  | | |  | |  |
| NMDP | 0.409 | 0.012 | | | 0.000 | | | 0.000 | | | 0.001 | | | 0.000 | | | 0.001 | | | 0.000 | | | 0.000 | | | 0.002 | | | | 0.000 | | | 0.000 | | | 0.000 | | | 0.000 | |  |
| Study | 0.501 | 0.037 | | | 0.003 | | | 0.000 | | | 0.005 | | | 0.002 | | | 0.011 | | | 0.000 | | | 0.000 | | | 0.012 | | | | 0.002 | | | 0.002 | | | 0.000 | | | 0.000 | |  |
| Black |  |  | | |  | | |  | | |  | | |  | | |  | | |  | | |  | | |  | | | |  | | |  | | |  | | |  | |  |
| NMDP | 0.028 | 0.009 | | | 0.365 | | | 0.000 | | | 0.000 | | | 0.000 | | | 1.389 | | | 0.000 | | | 0.000 | | | 1.023 | | | | 0.000 | | | 0.010 | | | 0.039 | | | 0.001 | |  |
| Study | 0.035 | 0.009 | | | 0.295 | | | 0.000 | | | 0.000 | | | 0.000 | | | 1.522 | | | 0.000 | | | 0.000 | | | 1.092 | | | | 0.000 | | | 0.007 | | | 0.050 | | | 0.007 | |  |
| Hispanic/Latinx |  |  | | |  | | |  | | |  | | |  | | |  | | |  | | |  | | |  | | | |  | | |  | | |  | | |  | |  |
| NMDP | 0.536 | 0.024 | | | 0.006 | | | 0.000 | | | 0.001 | | | 0.038 | | | 0.030 | | | 0.000 | | | 0.000 | | | 0.043 | | | | 0.000 | | | 0.000 | | | 0.000 | | | 0.000 | |  |
| Study | 0.692 | 0.044 | | | 0.015 | | | 0.004 | | | 0.000 | | | 0.131 | | | 0.084 | | | 0.000 | | | 0.000 | | | 0.106 | | | | 0.000 | | | 0.000 | | | 0.000 | | | 0.000 | |  |
| Asian |  |  | | |  | | |  | | |  | | |  | | |  | | |  | | |  | | |  | | | |  | | |  | | |  | | |  | |  |
| NMDP | 1.993 | 0.000 | | | 0.000 | | | 0.278 | | | 0.000 | | | 0.032 | | | 0.000 | | | 0.001 | | | 0.001 | | | 0.016 | | | | 0.000 | | | 0.000 | | | 0.000 | | | 0.000 | |  |
| Study | 3.238 | 0.021 | | | 0.000 | | | 0.797 | | | 0.000 | | | 0.041 | | | 0.010 | | | 0.000 | | | 0.000 | | | 0.010 | | | | 0.000 | | | 0.000 | | | 0.000 | | | 0.000 | |  |
|  | HLA-DR Antigens | | | | | | | | | | | | | | | | | | | | | | | | | | | | | | | |  | | |  | | |  | |  |
|  | 1 | 2 | | | 3 | | | 4 | | | 6 | | | 7 | | | 8 | | | 9 | | | 10 | | | 5 | | | |  | | |  | | |  | | |  | |  |
| White |  |  | | |  | | |  | | |  | | |  | | |  | | |  | | |  | | |  | | | |  | | |  | | |  | | |  | |  |
| NMDP | 1.200 | 2.506 | | | 1.315 | | | 2.483 | | | 2.342 | | | 1.683 | | | 0.089 | | | 0.010 | | | 0.009 | | | 1.642 | | | |  | | |  | | |  | | |  | |  |
| Study | 1.334 | 2.233 | | | 2.091 | | | 3.277 | | | 2.259 | | | 1.481 | | | 0.106 | | | 0.006 | | | 0.031 | | | 1.923 | | | |  | | |  | | |  | | |  | |  |
| Black |  |  | | |  | | |  | | |  | | |  | | |  | | |  | | |  | | |  | | | |  | | |  | | |  | | |  | |  |
| NMDP | 0.467 | 2.657 | | | 1.771 | | | 0.262 | | | 3.807 | | | 1.023 | | | 0.428 | | | 0.088 | | | 0.037 | | | 3.021 | | | |  | | |  | | |  | | |  | |  |
| Study | 0.386 | 2.757 | | | 2.004 | | | 0.254 | | | 3.745 | | | 0.977 | | | 0.495 | | | 0.113 | | | 0.046 | | | 3.237 | | | |  | | |  | | |  | | |  | |  |
| Hispanic/Latinx |  |  | | |  | | |  | | |  | | |  | | |  | | |  | | |  | | |  | | | |  | | |  | | |  | | |  | |  |
| NMDP | 0.745 | 1.325 | | | 0.675 | | | 4.394 | | | 3.159 | | | 1.156 | | | 0.865 | | | 0.014 | | | 0.026 | | | 1.016 | | | |  | | |  | | |  | | |  | |  |
| Study | 0.728 | 1.271 | | | 0.732 | | | 6.276 | | | 3.084 | | | 0.947 | | | 1.656 | | | 0.029 | | | 0.058 | | | 0.998 | | | |  | | |  | | |  | | |  | |  |
| Asian |  |  | | |  | | |  | | |  | | |  | | |  | | |  | | |  | | |  | | | |  | | |  | | |  | | |  | |  |
| NMDP | 0.077 | 4.101 | | | 0.319 | | | 1.681 | | | 2.163 | | | 0.987 | | | 0.273 | | | 0.591 | | | 0.138 | | | 2.915 | | | |  | | |  | | |  | | |  | |  |
| Study | 0.166 | 8.411 | | | 0.579 | | | 1.852 | | | 2.183 | | | 0.972 | | | 0.445 | | | 0.714 | | | 0.269 | | | 4.883 | | | |  | | |  | | |  | | |  | |  |
|  | HLA-DQ Antigens | | | | | | | | | |  | | |  | | |  | | |  | | |  | | |  | | | |  | | |  | | |  | | |  | |  |
|  | 1 | 2 | | | 3 | | | 4 | | |  | | |  | | |  | | |  | | |  | | |  | | | |  | | |  | | |  | | |  | |  |
| White |  |  | | |  | | |  | | |  | | |  | | |  | | |  | | |  | | |  | | | |  | | |  | | |  | | |  | |  |
| NMDP | 17.076 | 4.488 | | | 12.016 | | | 0.080 | | |  | | |  | | |  | | |  | | |  | | |  | | | |  | | |  | | |  | | |  | |  |
| Study | 15.576 | 5.138 | | | 13.114 | | | 0.112 | | |  | | |  | | |  | | |  | | |  | | |  | | | |  | | |  | | |  | | |  | |  |
| Black |  |  | | |  | | |  | | |  | | |  | | |  | | |  | | |  | | |  | | | |  | | |  | | |  | | |  | |  |
| NMDP | 22.636 | 4.619 | | | 5.691 | | | 0.500 | | |  | | |  | | |  | | |  | | |  | | |  | | | |  | | |  | | |  | | |  | |  |
| Study | 22.136 | 5.049 | | | 5.566 | | | 0.593 | | |  | | |  | | |  | | |  | | |  | | |  | | | |  | | |  | | |  | | |  | |  |
| Hispanic/Latinx |  |  | | |  | | |  | | |  | | |  | | |  | | |  | | |  | | |  | | | |  | | |  | | |  | | |  | |  |
| NMDP | 9.759 | 3.063 | | | 16.859 | | | 1.040 | | |  | | |  | | |  | | |  | | |  | | |  | | | |  | | |  | | |  | | |  | |  |
| Study | 8.901 | 2.734 | | | 19.379 | | | 1.820 | | |  | | |  | | |  | | |  | | |  | | |  | | | |  | | |  | | |  | | |  | |  |
| Asian |  |  | | |  | | |  | | |  | | |  | | |  | | |  | | |  | | |  | | | |  | | |  | | |  | | |  | |  |
| NMDP | 19.678 | 1.697 | | | 13.259 | | | 0.384 | | |  | | |  | | |  | | |  | | |  | | |  | | | |  | | |  | | |  | | |  | |  |
| Study | 24.498 | 1.800 | | | 14.184 | | | 0.559 | | |  | | |  | | |  | | |  | | |  | | |  | | | |  | | |  | | |  | | |  | |  |

### **Supplementary Table 4.** Frequency of HLA Homozygosity in the Study Cohort Compared to the NMDP Registry

|  | HLA Locus | | | | |
| --- | --- | --- | --- | --- | --- |
|  | A | C | B | DR | DQ |
| White |  |  |  |  |  |
| NMDP | 16.775 | 15.968 | 8.133 | 13.278 | 33.660 |
| Study | 17.191 | 16.329 | 9.090 | 14.742 | 33.941 |
| Black |  |  |  |  |  |
| NMDP | 17.962 | 12.977 | 8.119 | 14.013 | 33.344 |
| Study | 18.225 | 15.503 | 7.835 | 13.560 | 33.446 |
| Hispanic/Latinx |  |  |  |  |  |
| NMDP | 17.218 | 13.315 | 7.681 | 13.374 | 30.720 |
| Study | 18.796 | 13.845 | 9.254 | 15.778 | 32.835 |
| Asian |  |  |  |  |  |
| NMDP | 17.209 | 14.061 | 8.219 | 13.247 | 35.018 |
| Study | 23.019 | 17.794 | 12.818 | 20.474 | 41.041 |

**Supplementary Figure 2.** Plot of CPRA Sensitization by Total Number of Homozygous Loci

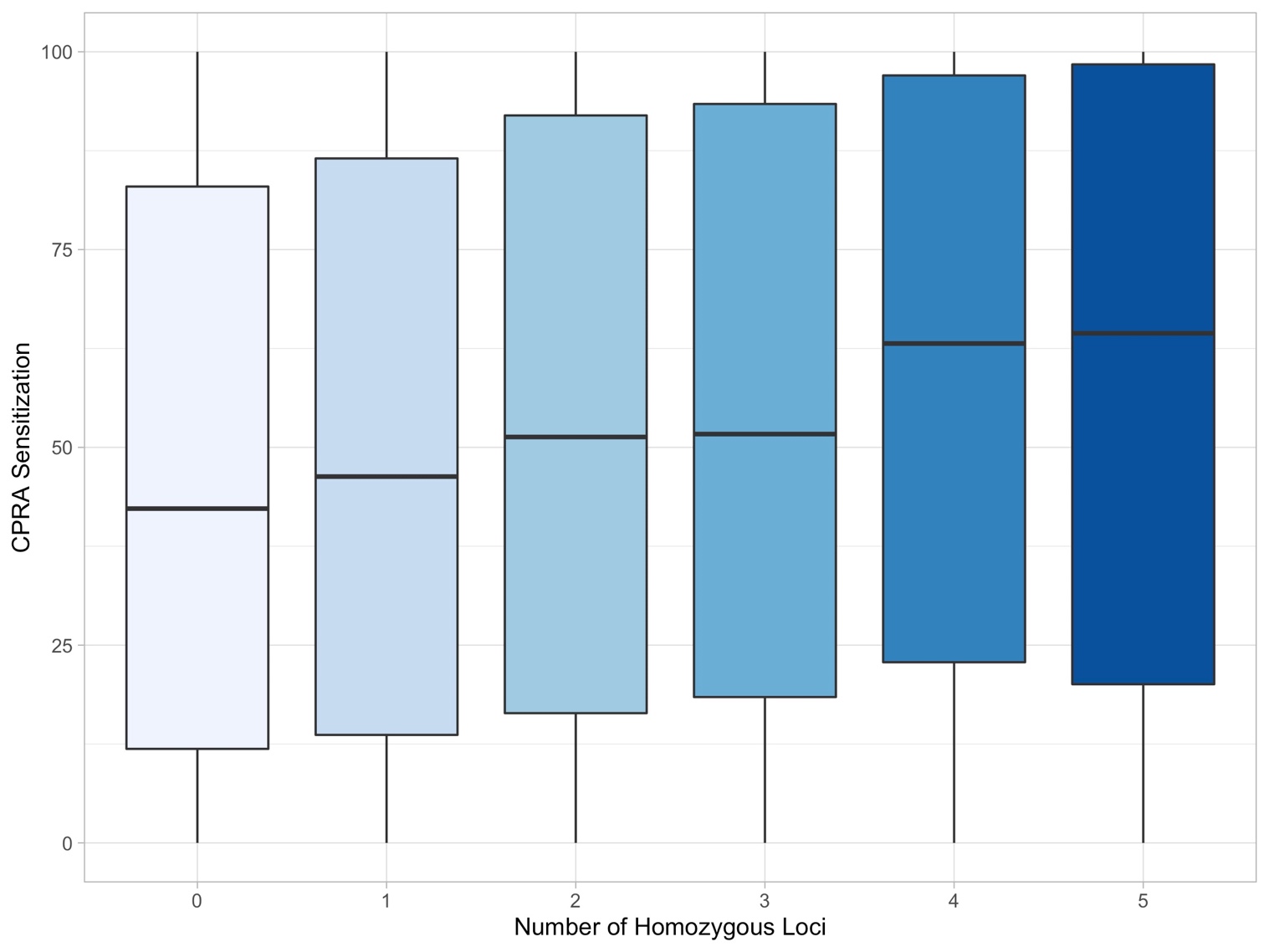

### **Supplementary Table 5**. Odds of Presence in Sensitization Groups Based on Homozygosity at a Single HLA Locus

|  | Mildly-Sensitized  (CPRA 1-69) |  | Highly-Sensitized  (CPRA 70-94) |  | Extremely-Sensitized  (CPRA 95-100) |  |
| --- | --- | --- | --- | --- | --- | --- |
|  | OR (95% CI) | p-value | OR (95% CI) | p-value | OR (95% CI) | p-value |
| White | | | | | | |
| HLA - A | 1.076 (1.025-1.128) | 0.003 | 1.065 (0.982 -1.156) | 0.128 | 1.314 (1.215-1.421) | <0.001 |
| HLA - C | 1.166 (1.111 – 1.225) | <0.001 | 1.249 (1.153 – 1.345) | <0.001 | 1.546 (1.431 – 1.671) | <0.001 |
| HLA - B | 1.164 (1.093- 1.239) | <0.001 | 1.171 (1.054 – 1.301) | 0.003 | 1.765 (1.607 – 1.939) | <0.001 |
| HLA - DR | 1.042 (0.989 – 1.097) | 0.121 | 1.067 (0.978 – 1.164) | 0.146 | 1.898 (1.759 – 2.048) | <0.001 |
| HLA - DQ | 1.046 (1.006 – 1.087) | 0.022 | 1.200 (1.126 – 1.280) | <0.001 | 1.553 (1.456 – 1.655) | <0.001 |
| Black | | | | | | |
| HLA - A | 1.048 (0.991 – 1.108) | 0.100 | 1.093 (1.003 – 1.192) | 0.044 | 1.322 (1.228 – 1.423) | <0.001 |
| HLA - C | 1.032 (0.968 -1.099) | 0.337 | 0.979 (0.885 – 1.084) | 0.685 | 1.272 (1.170 – 1.384) | <0.001 |
| HLA - B | 1.081 (1.000 – 1.170) | 0.051 | 1.121 (0.993 – 1.266) | 0.066 | 1.430 (1.293 – 1.582) | <0.001 |
| HLA - DR | 1.087 (1.022 - 1.156) | 0.008 | 1.124 (1.021 – 1.237) | 0.017 | 1.455 (1.344 – 1.577) | <0.001 |
| HLA - DQ | 1.044 (0.998 – 1.093) | 0.060 | 1.120 (1.043 – 1.202) | 0.002 | 1.337 (1.257 - 1.423) | <0.001 |
| Hispanic/Latinx | | | | | | |
| HLA - A | 1.053 (0.980 – 1.132) | 0.160 | 1.025 (0.906 – 1.159) | 0.696 | 1.181 (1.055 – 1.323) | 0.004 |
| HLA - C | 1.162 (1.071 – 1.261) | <0.001 | 1.294 (1.134 – 1.478) | <0.001 | 1.524 (1.351 – 1.719) | <0.001 |
| HLA - B | 1.057 (0.958 – 1.167) | 0.271 | 1.249 (1.067 – 1.462) | 0.006 | 1.480 (1.284 – 1.705) | <0.001 |
| HLA - DR | 1.041 (0.962 – 1.126) | 0.317 | 1.141 (1.002 – 1.299) | 0.047 | 1.768 (1.583 – 1.974) | <0.001 |
| HLA - DQ | 1.004 (0.945 - 1.067) | 0.243 | 1.063 (0.960 – 1.177) | 0.243 | 1.402 (1.277 – 1.541) | <0.001 |
| Asian | | | | | | |
| HLA - A | 1.157 (1.036 – 1.291) | 0.010 | 1.141 (0.946 – 1.376) | 0.169 | 1.309 (1.094 – 1.566) | 0.003 |
| HLA - C | 1.014 (0.896 – 1.147) | 0.830 | 1.433 (1.181 – 1.738) | <0.001 | 1.470 (1.217 – 1.776) | <0.001 |
| HLA - B | 1.059 (0.919 – 1.221) | 0.426 | 1.179 (0.933 – 1.490) | 0.167 | 1.935 (1.585 – 2.364) | <0.001 |
| HLA - DR | 1.113 (0.991 – 1.250) | 0.071 | 1.359 (1.126 – 1.642) | 0.001 | 1.687 (1.413 – 2.015) | <0.001 |
| HLA - DQ | 1.278 (1.163 – 1.405) | <0.001 | 1.330 (1.132 – 1.563) | 0.001 | 1.525 (1.302- 1.787) | <0.001 |

### **Supplementary Table 6**. Odds of Presence in Sensitization Groups Based on Homozygosity at Multiple HLA Loci

|  | Mildly-Sensitized  (CPRA 1-69) |  | Highly-Sensitized  (CPRA 70-94) |  | Extremely-Sensitized  (CPRA 95-100) |  |
| --- | --- | --- | --- | --- | --- | --- |
|  | OR (95% CI) | p-value | OR (95% CI) | p-value | OR (95% CI) | p-value |
| White | | | | | | |
| 1 Locus | 1.066 (1.026 - 1.112) | 0.003 | 1.164 (1.083 - 1.251) | <0.001 | 1.342 (1.242 - 1.450) | <0.001 |
| 2 Loci | 1.088 (1.032 - 1.148) | 0.002 | 1.221 (1.117 - 1.336) | <0.001 | 1.833 (1.678 – 2.003) | <0.001 |
| 3 Loci | 1.191 (1.093 - 1.298) | <0.001 | 1.299 (1.126 – 1.499) | <0.001 | 2.226 (1.955 - 2.535) | <0.001 |
| 4 or 5 Loci | 1.276 (1.144 - 1.424) | <0.001 | 1.482 (1.243 – 1.768) | <0.001 | 3.331 (2.885 – 3.847) | <0.001 |
| Black | | | | | | |
| 1 Locus | 1.010 (0.962 - 1.060) | 0.691 | 1.028 (0.952 - 1.109) | 0.485 | 1.196 (1.115 -1.283) | <0.001 |
| 2 Loci | 1.098 (1.032 - 1.168) | 0.003 | 1.166 (1.059 - 1.284) | 0.002 | 1.627 (1.496 - 1.769) | <0.001 |
| 3 Loci | 1.232 (1.104 - 1.374) | <0.001 | 1.300 (1.100 - 1.537) | 0.002 | 2.043 (1.782 - 2.342) | <0.001 |
| 4 or 5 Loci | 0.971 (0.793 - 1.189) | 0.778 | 1.095 (0.804 - 1.491) | 0.565 | 2.341 (1.876- 2.922) | <0.001 |
| Hispanic/Latinx | | | | | | |
| 1 Locus | 1.036 (0.970 - 1.107) | 0.286 | 0.981 (0.875 – 1.099) | 0.737 | 1.167 (1.043 – 1.305) | 0.007 |
| 2 Loci | 1.036 (0.954 – 1.125) | 0.398 | 1.198 (1.047 – 1.370) | 0.009 | 1.654 (1.459 – 1.875) | <0.001 |
| 3 Loci | 1.206 (1.057 – 1.376) | 0.005 | 1.297 (1.045 – 1.611) | 0.018 | 1.731 (1.419 – 2.111) | <0.001 |
| 4 or 5 Loci | 1.148 (0.948 – 1.391) | 0.157 | 1.340 (0.988 – 1.817) | 0.060 | 2.992 (2.379– 3. 627) | <0.001 |
| Asian | | | | | | |
| 1 Locus | 1.231 (1.098 – 1.380) | <0.001 | 1.215 (0.994 – 1.468) | 0.057 | 1.310 (1.065 – 1.612) | 0.011 |
| 2 Loci | 1.382 (1.214 – 1.573) | <0.001 | 1.382 (1.104 – 1.730) | 0.005 | 1.750 (1.402 – 2.184) | <0.001 |
| 3 Loci | 1.147 (0.952 – 1.382) | 0.149 | 1.469 (1.089 – 1.980) | 0.012 | 1.941 (1.459 – 2.582) | <0.001 |
| 4 or 5 Loci | 1.419 (1.113 -1.809) | 0.005 | 2.148 (1.503 – 3.070) | <0.001 | 3.493 (2.542 -4.801) | <0.001 |

### **Supplementary Table 7.** Single Variable Modeling of Factors Associated with Sensitization

|  | Sensitization Group | | | | | |
| --- | --- | --- | --- | --- | --- | --- |
|  | Mildly-sensitized  (CPRA 1-69) | | Highly-sensitized  (CPRA 70-94) | | Extremely-sensitized  (CPRA 95-100) | |
|  | OR (95% CI) | p-value | OR (95% CI) | p-value | OR (95% CI) | p-value |
| Number Homozygous HLA Loci | 1.05 (1.03-1.06) | <0.001 | 1.08 (1.04-1.11) | <0.001 | 1.27 (1.25-1.29) | <0.001 |
| Sex |  |  |  |  |  |  |
| Male | reference |  | reference |  | reference |  |
| Female | 1.54 (1.50-1.58) | <0.001 | 4.76 (4.57-4.97) | <0.001 | 4.09 (3.93-4.25) | <0.001 |
| Sex * HLA Homozygosity |  |  |  |  |  |  |
| Male | 0.98 (0.97-0.99) | 0.006 | 0.72 (0.70-0.75) | <0.001 | 0.90 (0.88-0.92) | <0.001 |
| Female | 1.21 (1.19-1.23) | <0.001 | 1.60 (1.56-1.63) | <0.001 | 1.82 (1.79-1.86) | <0.001 |
| Age (per decade) | 0.98 (0.97-0.99) | <0.001 | 0.96 (0.94-0.97) | <0.001 | 0.85 (0.84-0.86) | <0.001 |
| Age * HLA Homozygosity | 1.01 (1.00-1.01) | <0.001 | 1.01 (1.01-1.02) | <0.001 | 1.03 (1.03-1.04) | <0.001 |
| Ethnicity |  |  |  |  |  |  |
| White | reference |  | reference |  | reference |  |
| Black | 1.36 (1.33-1.40) | <0.001 | 1.50 (1.43-1.57) | <0.001 | 2.05 (1.97-2.14) | <0.001 |
| Hispanic/Latinx | 0.93 (0.89-0.96) | <0.001 | 0.94 (0.89-1.00) | 0.036 | 1.10 (1.04-1.176) | 0.001 |
| Asian | 1.04 (0.99-1.09) | 0.153 | 1.01 (0.93-1.10) | 0.841 | 1.09 (1.00-1.19) | 0.042 |
| Ethnicity * HLA Homozygosity |  |  |  |  |  |  |
| White | 1.01 (1.00-1.03) | 0.081 | 1.04 (1.01-1.06) | 0.005 | 1.16 (1.13-1.18) | <0.001 |
| Black | 1.19 (1.17-1.21) | <0.001 | 1.26 (1.23-1.30) | <0.001 | 1.61 (1.57-1.65) | <0.001 |
| Hispanic/Latinx | 0.97 (0.94-0.99) | 0.002 | 1.00 (0.96-1.04) | 0.949 | 1.18 (1.14-1.21) | <0.001 |
| Asian | 1.04 (1.01-1.07) | 0.005 | 1.07 (1.02-1.12) | 0.005 | 1.17 (1.12-1.22) | <0.001 |
| Diabetes |  |  |  |  |  |  |
| None | reference |  | reference |  | reference |  |
| Type 1 | 0.97 (0.91-1.04) | 0.422 | 0.99 (0.88-1.10) | 0.785 | 1.18 (1.08-1.29) | <0.001 |
| Type 2 | 0.95 (0.93-0.97) | <0.001 | 0.75 (0.72-0.78) | <0.001 | 0.55 (0.52-0.57) | <0.001 |
| Unknown Type | 0.91 (0.79-1.04) | 0.151 | 0.90 (0.72-1.12) | 0.342 | 1.18 (0.98-1.41) | 0.075 |
| Diabetes * HLA Homozygosity |  |  |  |  |  |  |
| None | 1.06 (1.04-1.07) | <0.001 | 1.13 (1.10-1.15) | <0.001 | 1.36 (1.33-1.38) | <0.001 |
| Type 1 | 1.03 (0.98-1.08) | 0.229 | 1.13 (1.05-1.21) | 0.001 | 1.48 (1.40-1.56) | <0.001 |
| Type 2 | 1.03 (1.02-1.05) | <0.001 | 1.01 (0.98-1.04) | 0.609 | 1.05 (1.02-1.08) | <0.001 |
| Unknown Type | 0.95 (0.86-1.05) | 0.312 | 1.00 (0.85-1.17) | 0.995 | 1.37 (1.22-1.53) | <0.001 |
| Blood Type |  |  |  |  |  |  |
| O | reference |  | reference |  | reference |  |
| A | 0.93 (0.90-0.95) | <0.001 | 0.95 (0.90-0.99) | 0.015 | 0.98 (0.93-1.02) | 0.252 |
| B | 1.06 (1.03-1.10) | <0.001 | 1.02 (0.96-1.08) | 0.571 | 1.09 (1.03-1.15) | 0.002 |
| AB | 0.89 (0.84-0.95) | 0.001 | 0.99 (0.90-1.10) | 0.904 | 1.00 (0.90-1.10) | 0.976 |
| Blood Type * HLA Homozygosity |  |  |  |  |  |  |
| O | 1.06 (1.04-1.07) | <0.001 | 1.09 (1.06-1.11) | <0.001 | 1.26 (1.24-1.29) | <0.001 |
| A | 1.02 (1.00-1.04) | 0.045 | 1.07 (1.04-1.11) | <0.001 | 1.26 (1.23-1.29) | <0.001 |
| B | 1.08 (1.06-1.11) | <0.001 | 1.09 (1.05-1.14) | <0.001 | 1.29 (1.25-1.33) | <0.001 |
| AB | 0.99 (0.94-1.04) | 0.668 | 1.11 (1.03-1.19) | 0.005 | 1.27 (1.19-1.35) | <0.001 |
| Prior Kidney Txp |  |  |  |  |  |  |
| No | reference |  | reference |  | reference |  |
| Yes | 1.98 (1.88-2.07) | <0.001 | 8.73 (8.29-9.20) | <0.001 | 25.83 (24.62-27.10) | <0.001 |
| Prior Kidney Txp * HLA Homozygosity |  |  |  |  |  |  |
| No | 1.03 (1.02-1.05) | <0.001 | 0.96 (0.94-0.98) | <0.001 | 0.92 (0.90-0.94) | <0.001 |
| Yes | 1.57 (1.50-1.64) | <0.001 | 3.29 (3.15-3.44) | <0.001 | 4.90 (4.71-5.10) | <0.001 |

Txp = Transplant
